# Clinical and Therapeutic Applications of Individual-level Tissue-Specific Imputed Transcriptomes

**DOI:** 10.1101/2022.11.23.22282644

**Authors:** Xin Wang, Caitlin Selvaggi, Lu-Chen Weng, Sean J. Jurgens, Seung Hoan Choi, Anjali Jha, Tingyi Cao, Satoshi Koyama, Joel T. Rämö, Shinwan Kany, Patrick T. Ellinor, Andrea S. Foulkes, Steven A. Lubitz

**Affiliations:** Cardiovascular Research Center, Massachusetts General Hospital, Boston, MA, USA; Cardiovascular Disease Initiative, The Broad Institute of MIT and Harvard, Cambridge, MA, USA; Biostatistics Center, Massachusetts General Hospital, Boston, MA, USA; Department of Experimental Cardiology, Amsterdam Cardiovascular Sciences, Amsterdam UMC, Amsterdam, NL; Department of Biostatistics, Boston University, Boston, MA, USA; Geisel School of Medicine, Dartmouth College; Department of Biostatistics, Harvard T.H. Chan School of Public Health, Boston, MA, USA; Institute for Molecular Medicine Finland (FIMM), Helsinki Institute of Life Science (HiLIFE), University of Helsinki, Helsinki, Finland; Department of Cardiology, University Heart and Vascular Center Hamburg, Hamburg, Germany; Department of Medicine, Harvard Medical School, Boston, MA, USA

## Abstract

Gene expression causally contributes to phenotypic variability. However, since tissue-specific gene expression is difficult to measure at scale, inferring phenotypic risk for individuals using expression profiles can be difficult. We statistically imputed tissue-specific gene expression for 486,452 UK Biobank participants and demonstrated its applications using rich phenotypic data in the biorepository. We performed joint analysis of transcriptome-wide association studies (TWAS) and colocalization for six traits to prioritize tissue-gene pairs for therapeutics development. Using genes identified in TWAS, we constructed imputed transcriptome risk scores and showed their predictive utility even when conditioning on genome-wide polygenic risk scores. A phenome-wide association analysis for established therapeutic targets in whole blood recapitulated known biology and prioritized repurposing potentials supported by previous literature. Our study provides evidence supporting the utility of individual-level imputed transcriptomes for discovering novel gene-tissue-phenotype trios, expression-based phenotypic risk prediction, and identifying individuals with therapeutically targetable predisposition to disease.

## Introduction

Variability in gene expression influences phenotypic expression. However, gene expression is difficult to measure at scale due to tissue-specific effects and inaccessibility of various tissues. A class of methods referred to as transcriptome-wide association studies (TWAS) have emerged which are essentially gene-prioritization methods that associate genetically predicted gene expression levels with phenotypes.^1, 2^ In brief, such methods generally start with building statistical models that predict the *cis*genetic component of gene expression from genomic data using a dataset that comprise both reference transcriptome and genomic data, such as the Genotype-Tissue Expression (GTEx) project.^3^ The model weights can then be applied to an independent sample with genomic data to infer genetically regulated gene expression.

A popular application of TWAS is a method in which gene-trait associations are inferred using summary statistics from genome-wide association studies (GWAS) and pre-trained weights from models that predict expression from genetic variants.^4–8^ Through bypassing the step of imputing transcriptome at the individual level, this method does not require accessing genomic data to evaluate expression-phenotype associations and enables prioritization of disease-associated genes.^6^ In contrast to using GWAS summary level data, using individual-level imputed transcriptomes may have several specific attributes. For example, genetically predicted gene expression profiles may enable the identification of individuals who are most likely to express or develop a phenotype through specific gene expression pathways, some of which may be therapeutically targetable. Imputation of individual-level transcriptomes may also facilitate the scalable assessment of associations between predicted gene expression and a variety of phenotypes, rather than be limited to single trait-expression associations determined by availability of individual GWAS results. Moreover, since GWAS summary-level TWAS requires external reference panels to infer linkage disequilibrium (LD) for the GWAS cohort, the test statistics can be inaccurate if the reference population is dissimilar from the GWAS cohort.^1^

Large-scale biobanks with deep phenotypic and genomic data now enable the assessment and application of individual-level, tissue-specific, gene expression predictions. By imputing transcriptomes using genomic data, the prediction can serve as an intermediate molecular phenotype for biological discovery or be associated with multiple phenotypes within the same dataset to perform scalable TWAS. Here, we applied a joint-tissue imputation approach^8^ to the UK Biobank (UKBB) to predict genetically regulated gene expression levels across seven tissues in 486,452 participants. We assess the potential clinical and therapeutic applications of individual-level imputed transcriptomes in the context of large-scale biobanks using three diseases (atrial fibrillation [AF], coronary artery disease [CAD], type 2 diabetes [T2D]), three quantitative traits (heart rate corrected QT interval [QTc] on electrocardiograms [ECG], low-density lipoprotein cholesterol levels [LDL-C], glycated hemoglobin [HbA1c]), and 541 binary phenotypes created using International Classification of Diseases (ICD)-10 codes with the Phecode^9,10^ mapping system.

We first imputed the *cis* genetic component of expression for protein-coding genes in tissues that were biologically relevant to the curated phenotypes. We then performed a TWAS for each tissue-phenotype pair and processed the results using a locus-level colocalization analysis.^11^ We also examined the therapeutic targetability of prioritized genes to probe the potential both for drug repurposing and novel target discovery. Using suggestive genes obtained from the TWAS, we constructed two imputed transcriptome risk scores (ITRS) for each phenotype with all identified genes and a subset of druggable genes to predict risk in a biologically interpretable manner, and contrasted the approach with a conventional polygenic risk score (PRS) approach. Finally, we conducted phenome-wide association studies (PheWAS) for 541 phecodes (groups of similar ICD codes)^9, 10^ and 94 well-established therapeutically targetable genes expressed in the whole blood, to explore the potential for gene expression to guide identification of novel disease indications.

## Results

### Sample characteristics

The mean age of the study population (N=486,452) was 57.0 +/- 8.1 years at enrollment, and 54.2% (263,800) were female. Using principal components of ancestry, 426,759 (87.7%) participants were identified as being of White British ancestry. We identified 31,236 (6.4%) AF cases (8,287 prevalent cases at enrollment, 22,949 incident events during follow-up), 29,250 (6.0%) CAD cases (14,852 prevalent cases, 14,398 incident events), and 36,688 (7.5%) T2D cases (11,971 prevalent cases, 24,717 incident events; see **Methods** for phenotyping algorithms). Heart rate-corrected QT intervals on ECGs were calculated from resting 12-lead ECGs using the Bazett formula, with a sample size of 41,490 (mean value: 421.3 +/- 25.7 ms). LDL-C and HbA1c levels were extracted from biochemistry assay measurements collected at enrollment. 462,739 individuals had LDL-C data available (mean value: 3.6 +/- 0.9 mmol/L) and 462,159 had HbA1c data available (mean value: 36.13 +/- 6.77 mmol/mol). Descriptive statistics are presented in **Table 1**.

**Table 1.** Sample characteristics.

|  | **N=486,452**<br>**Mean (SD) or N (%)** |
| --- | --- |
| **Age at enrollment** | 57.04 (8.09) |
| **Female sex** | 263,800 (54.23%) |
| **White British ancestry** | 426,759 (87.73%) |
| **Atrial fibrillation** | 31,236 (6.42%) |
| Prevalent disease | 8,287 (1.70%) |
| Incident disease | 22,949 (4.72%) |
| **Coronary artery disease** | 29,250 (6.01%) |
| Prevalent disease | 14,852 (3.05%) |
| Incident disease | 14,398 (2.96%) |
| **Type 2 diabetes** | 36,688 (7.54%) |
| Prevalent disease | 11,971 (2.46%) |
| Incident disease | 24,717 (5.08%) |
|  | **N=41,490** |
| **QTc interval on electrocardiograms (ms)** | 421.31 (25.70) |
|  | **N=462,739** |
| **Low-density lipoprotein cholesterol (mmol/L)** | 3.56 (0.87) |
|  | **N=462,159** |
| **Glycated hemoglobin (mmol/mol)** | 36.13 (6.77) |

**White British ancestry:** determined using principal components of ancestry
**QTc interval:** heart rate-corrected QT interval on electrocardiograms

Using ICD-10 codes from UKBB data, we derived 1,590 phecodes^9, 10^ with non-zero cases, of which 629 represented independent index phecodes using a hierarchical clustering algorithm (see **Methods**). The mapping algorithm takes ICD codes as input and automatically excludes non-eligible controls for each phecode (e.g., subjects with type 1 diabetes are excluded from the type 2 diabetes phecode control set), and accounts for sex-specific phenotypes (e.g., menopausal and postmenopausal disorders). Thus phecode datasets vary in sample size. We restricted the phecodes to those with greater than 20 cases, resulting in 541 phecodes for analysis. The mean phecode cases and sample sizes are 1,887 cases (minimum 21; maximum 95,961) and 447,789 samples (minimum 216,628; maximum 486,366), respectively.

### Trait-relevant tissues and transcriptome imputation

For each of the six curated phenotypes, we selected biologically relevant tissues available from GTEx *a priori*, including for AF (right atrial appendage and left ventricle), CAD (subcutaneous adipose, liver, and coronary artery), T2D (subcutaneous adipose, liver, and pancreas), QTc interval (left ventricle), LDL-C (liver), and HbA1c (subcutaneous adipose, liver, and pancreas). For each tissue, we predicted the *cis* genetic component of gene expression for all imputable protein-coding genes on autosomes using a joint-tissue imputation approach,^8^ resulting in 9,747, 9,077, 10,531, 6,373, 7,745, and 8,689 genes in the atrial appendage, left ventricle, subcutaneous adipose, liver, coronary artery, and pancreas tissue, respectively. This method trained expression prediction models using the residuals of the normalized expression levels after adjusting for covariates, and included as predictors biallelic single nucleotide polymorphism (SNPs) within 1 Mb of the predicted gene.^8^ Imputable genes were defined as genes with available prediction models; gene information can be found in the GENCODE v37 annotation dataset.^12^ Across all imputable protein-coding genes in the identified tissues, the mean number of SNPs included in a prediction model for a single gene was 12.9 (standard deviation: 10.1).^8^

### TWAS, colocalization, and druggability of prioritized genes

The study overview is shown in **Figure 1**. We tested associations between imputed expression of protein-coding genes and curated phenotypes in their biologically relevant tissues using generalized regression models in 486,452 UK Biobank participants. For each model, the phenotype was regressed on the genetically predicted expression of a single gene (rank-based inverse normal transformed), with adjustment for enrollment age, sex, genotyping platform, and the top 5 principal components of ancestry. We identified 1,082 gene-tissue-phenotype trios (e.g., the *cis* genetic component of *ANGPTL3* expression in the liver was positively associated with LDL-C levels) at a Bonferroni corrected significance *P* value < 4.5×10^-7^ (0.05/110,109 tests) and 2,973 gene-tissue-phenotype trios at a suggestive significance level defined as a false discovery rate adjusted *P* value < 0.01.

**Figure 1.**
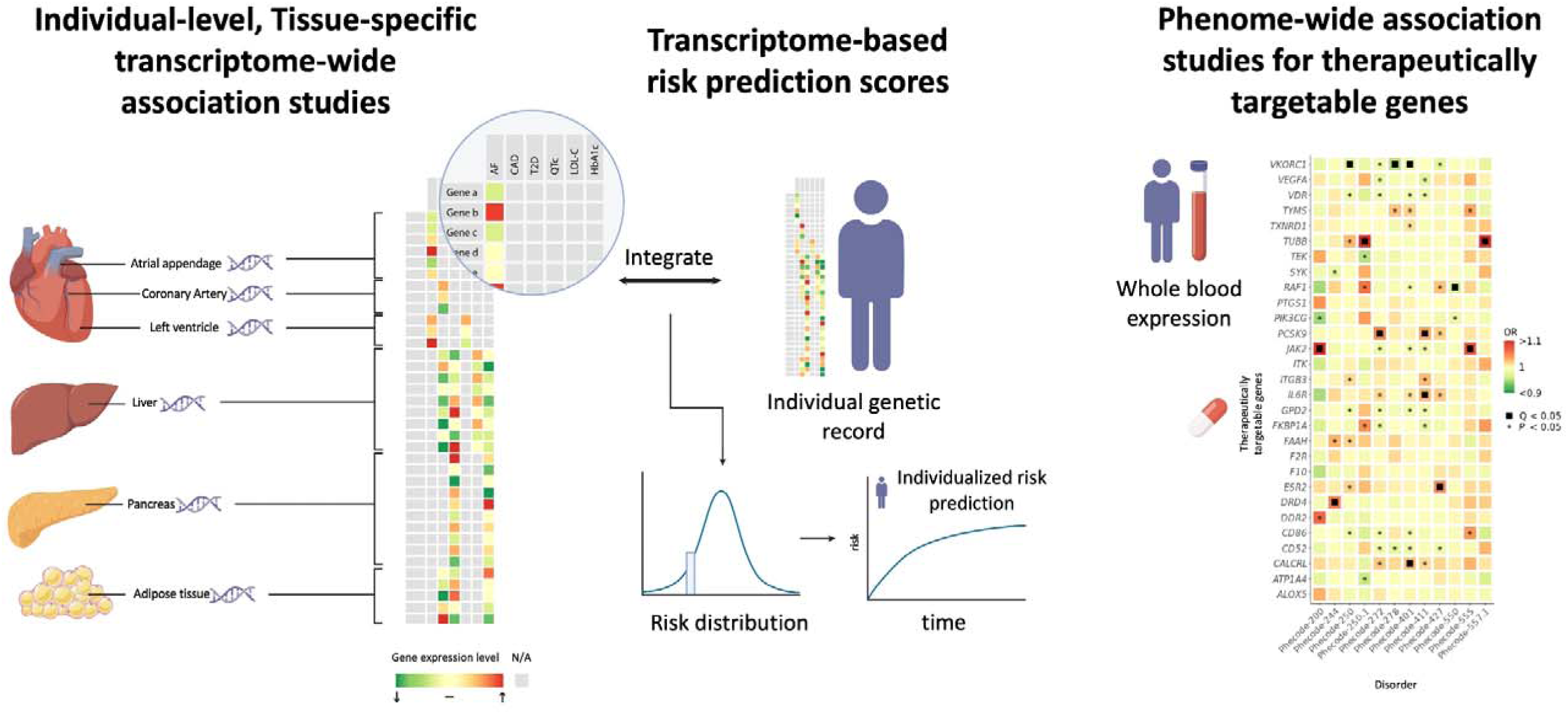
Study overview. **AF:** atrial fibrillation; **CAD:** coronary artery disease; **T2D:** type 2 diabetes; **QTc:** heart rate-corrected QT interval on electrocardiograms; **LDL-C:** low-density lipoprotein cholesterol; **HbA1c:**glycated hemoglobin; **Phecodes:** disease phenotypes created by grouping similar ICD-10 codes.^10^

Since TWAS signals tend to cluster around significant trait-associated SNPs due to linkage disequilibrium (LD), and the causal expression quantitative trait loci (eQTL) and the lead SNPs do not need to be colocalized to drive a TWAS signal,^11, 13^ we filtered the Bonferroni-significant TWAS results using a locus-level colocalization analysis^11^ to prioritize high-confidence signals. Among the 1,082 identified gene-tissue-phenotype trios, 144 were further supported by colocalization analysis using a gene-level colocalization probability [GLCP] threshold of > 0.50 (**Supplemental Table 1**). Summary statistics of all tests are uploaded as a **Supplemental Data File**.

We annotated therapeutic targetability for the 144 gene-tissue-phenotype trios prioritized by the joint analysis using a dataset by Gaziano et al,^14^ which contains actionable human proteins that are drug targets of approved or clinical-trial-stage medications identified using the ChEMBL v.26 database.^15^ Among the signals supported by TWAS and colocalization analysis, we identified 12 gene-tissue-phenotype trios for which the genes were potential therapeutic targets (**Figure 2**), including 4 trios for which the genes were targets of approved medications and the effect of medications were consistent with the direction of lowering phenotypic risk. Using the Open Targets Platform^16^ to retrieve indications for these medications, we observed one signal for which the tested phenotype was consistent with the approved indication, while the remaining signals reflect therapeutic targets for other indications (**Table 2**).

**Figure 2.**
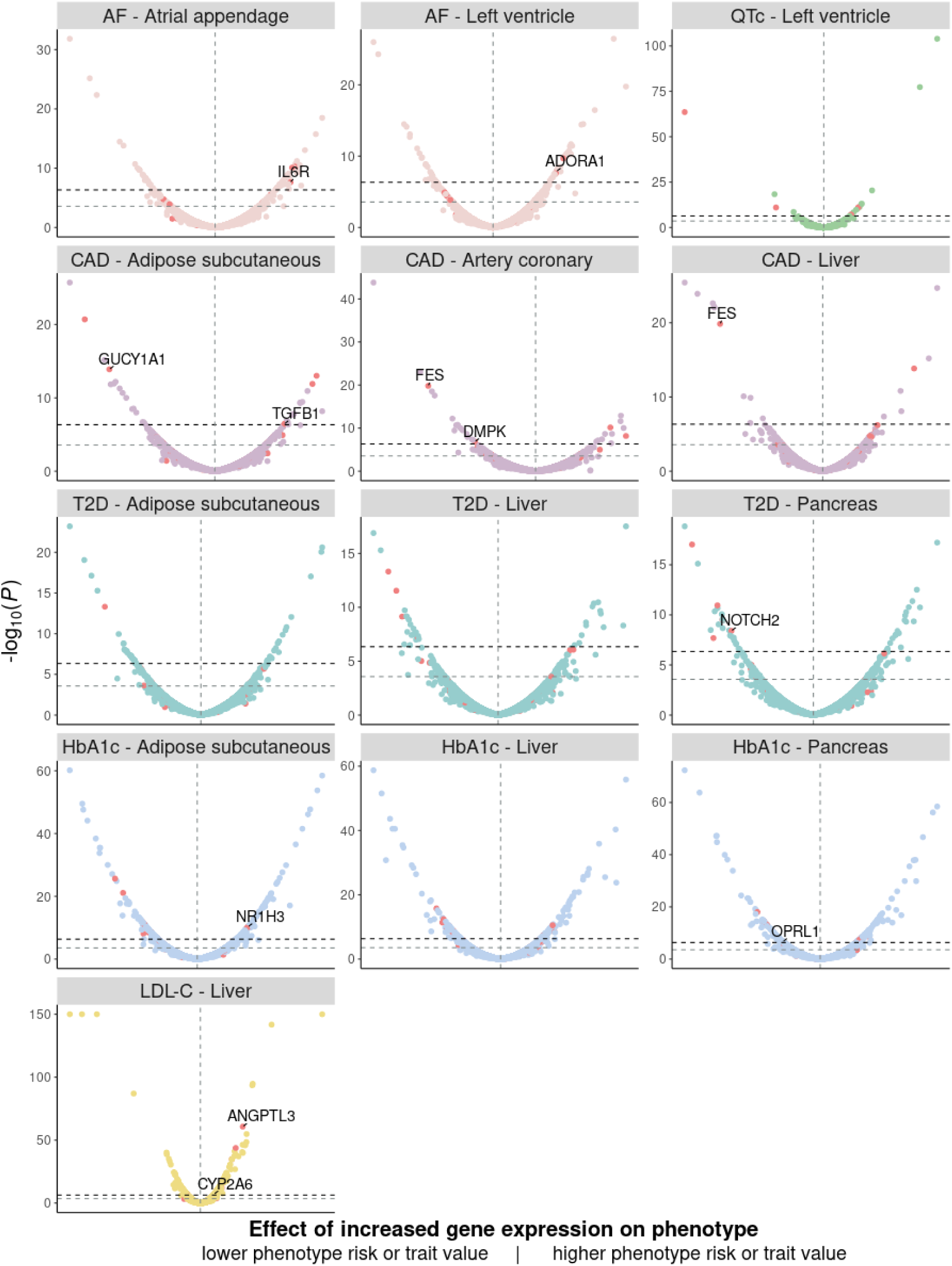
Transcriptome-wide association studies and druggability of prioritized trait-associated genes. **AF:** atrial fibrillation; **QTc:** heart rate-corrected QT interval on electrocardiograms; **CAD:** coronary artery disease; **T2D:**type 2 diabetes; **HbA1c:** glycated hemoglobin; **LDL-C:** low-density lipoprotein cholesterol. Effect sizes of tested gene expression levels on traits are plotted on the x-axis (log odds for disease phenotypes, beta estimates for quantitative traits). The vertical dashed line on each plot represents zero effect size. Genes on the right side are positively associated with phenotypic risk. The -log10 (*P* values) of association tests are plotted on the y-axis. The significance threshold determined by Bonferroni correction (*P* < 0.05/110,109 tests) is indicated by the dark horizontal dashed line, and the suggestive significance threshold determined by false discovery rate (adjusted *P* value < 0.01) is indicated by the gray horizontal dashed line. Figure titles specify phenotype-tissue pairs (a phenotype with one of its biologically relevant tissues). Each dot represents an association between the phenotype and the genetically predicted expression of a protein-coding gene in that tissue. Druggable genes (see text for definition) are plotted as red dots in each panel. Associations that passed the significance threshold, were further supported by colocalization analysis, and involved a druggable gene are annotated by gene names.

**Table 2.** Prioritized TWAS signals involving established therapeutic targets.

| Gene | Tissue | Tested phenotype | Effect size [95% CI] | P value (TWAS) | GLCP | Medication (Mechanism) | Indications |
| --- | --- | --- | --- | --- | --- | --- | --- |
| *ANGPTL3\** | Liver | LDL-C | 0.02 [0.02, 0.02] | 1.79E-61 | 0.95 | Evinacumab (inhibitor) | Familial hypercholesterolemia |
| *GUCY1A1* | Subcutaneous adipose | CAD | 0.95 [0.94, 0.96] | 1.28E-14 | 0.98 | Vericiguat (activator) | Heart failure |
| *IL6R* | Atrial appendage | AF | 1.04 [1.02, 1.05] | 2.41E-8 | 0.80 | Tocilizumab (inhibitor) | Rheumatoid arthritis, systemic juvenile idiopathic arthritis, systemic sclerosis, giant cell arteritis |
| *TGFB1* | Subcutaneous adipose | CAD | 1.03 [1.02, 1.04] | 3.67E-7 | 1.00 | Luspatercept (inhibitor) | Anemia, beta thalassemia, myelodysplastic syndromes |

**TWAS:** transcriptome-wide association studies; **LDL-C:** low-density lipoprotein cholesterol; **CAD:** coronary artery disease; **AF:** atrial fibrillation; **Effect size:** odds ratios for AF and CAD, beta estimate for LDL-C; corresponding to per 1 standard deviation increase in rank-based inverse normal transformed imputed gene expression levels; **95% CI:** 95% confidence interval of the effect size; **GLCP:** gene-level colocalization probability. **Medication (Mechanism):** approved medication targeting the tested gene and its mechanism; **Indications:** reported indications retrieved from the Open Targets Platform.<sup>16</sup>
\*Association for which the tested phenotype was consistent with the indication.

### Predictive utility of tissue-informed imputed transcriptome risk scores

For each curated phenotype, we constructed two imputed transcriptome risk scores (ITRS) comprising imputable protein-coding genes identified at the suggestive significance level from the phenotype-specific TWAS. The first score (ITRS-transcriptome) comprised all associated genes, and the second (ITRS-drug) comprised the subset of genes encoding potential therapeutic targets. For each score, gene-tissue pairs (gene expression levels in a specific tissue) identified for the corresponding phenotype were put into an elastic net model in a derivation dataset to select features and adjust weights. Scoring weights were then applied to a validation dataset to calculate ITRS and evaluate their predictive utility (the derivation-validation framework is shown in **Supplemental Figure1**). Gene-tissue pairs and their weights for ITRS-drug and ITRS-transcriptome are summarized in **Supplemental Table 2** and **Supplemental Table 3**, respectively. We observed significant associations between all ITRS-transcriptome and their corresponding phenotypes in validation sets, with hazard ratios (HRs) ranging from 1.30 to 1.39 per standard deviation (SD) increase in the score for disease phenotypes and beta estimates ranging from 0.15 to 4.98 (depending on trait units) per SD increase in the score for quantitative traits. We observed persistent associations with attenuated effect sizes when testing the associations between ITRS-drug and the phenotypes, with HRs ranging from 1.09 to 1.13 for diseases and betas ranging from 0.04 to 2.39 for quantitative traits (**Figure 3, Supplemental Table 4**).

**Figure 3.**
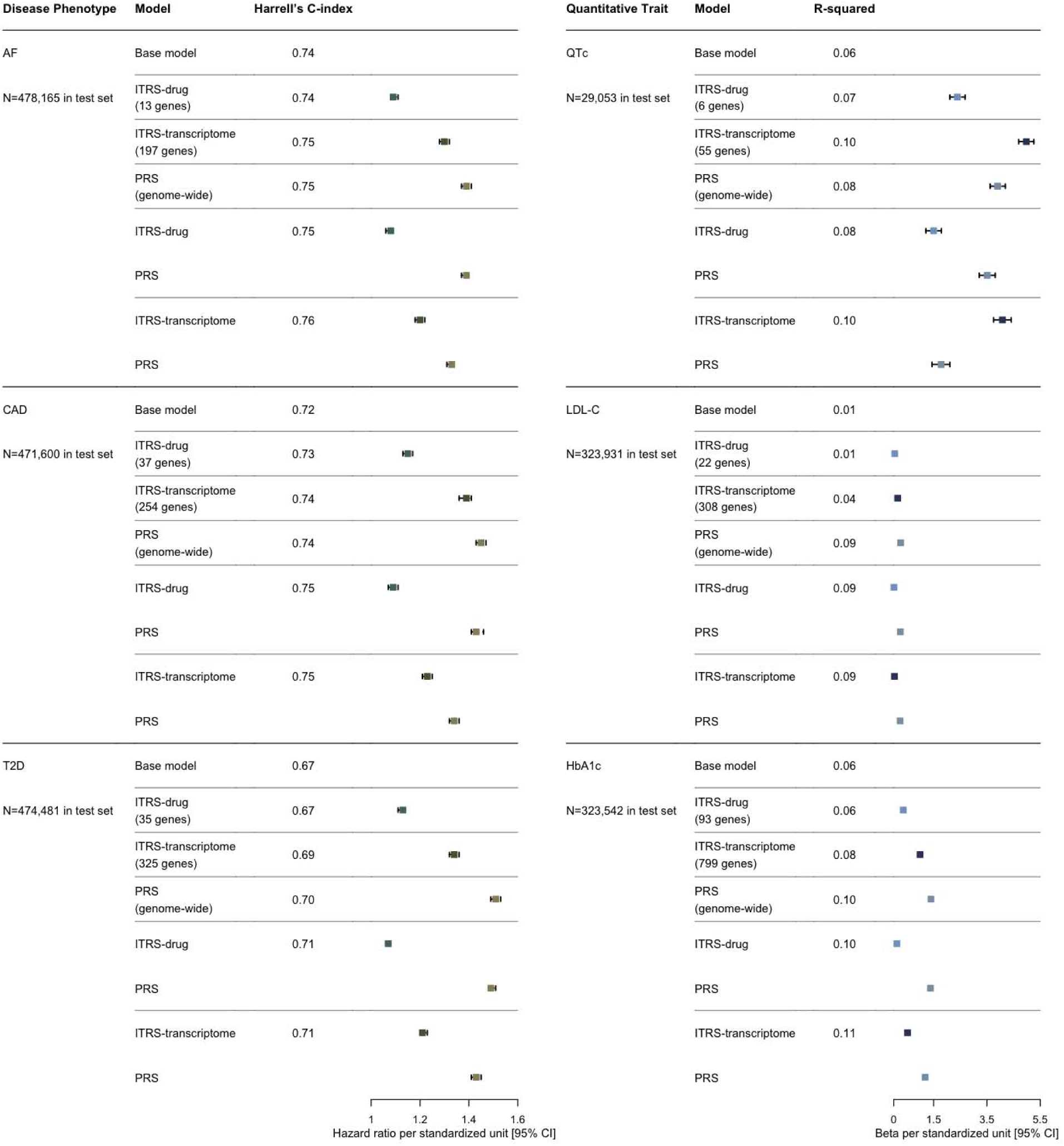
Predictive utility of imputed transcriptome risk scores and polygenic risk scores for select phenotypes. Each row represents a model. Risk scores were first tested individually in a model for association with the corresponding phenotype, adjusting for covariates. The imputed transcriptome risk scores (ITRS) for each phenotype were then tested in separate models together with corresponding genome-wide polygenic risk scores (PRS), adjusting for the same covariates. **AF:** atrial fibrillation; **CAD:**coronary artery disease; **T2D:** type 2 diabetes; **QTc:** heart rate-corrected QT interval on electrocardiograms; **LDL-C:** low-density lipoprotein cholesterol; **HbA1c:** hemoglobin A1C (glycated hemoglobin); **Base model:** models that only contain covariates (age, sex, genotyping platform, top 5 principal components of ancestry [PCs]); **ITRS-transcriptome:** imputed transcriptome risk score constructed using suggestive significant tissue-gene pairs observed in transcriptome-wide association study (TWAS); **ITRS-drug:** ITRS constructed using a subset of suggestive significant tissue-gene pairs observed in TWAS, for which the genes are actionable proteins defined in a previous study;^14^ **PRS:** genome-wide polygenic risk score; **Harrell’s C-index:** Harrell’s concordance index of the model; **R-squared**: variation in traits explained by variables in the model; **Hazard ratio:** hazard ratio in Cox proportional hazard models, corresponding to 1 standard deviation (SD) increase in risk scores; **95% CI:** 95% confidence intervals; **Beta:** effect size estimates in linear regression models, corresponding to 1 SD increase in risk scores.

To compare the predictive value of ITRS to that of a more conventional genetic risk prediction approach using polygenic risk scores (PRS), we constructed a PRS for each phenotype using PRS-CS-auto^17^ and tested its association with the trait using the same approach. The scoring weights for calculating PRS are uploaded as **Supplemental Data Files**. As expected, we observed significant associations between each PRS and its corresponding phenotype, with HRs ranging from 1.39 to 1.51 per SD increase in the score for disease phenotypes and betas ranging from 0.26 to 3.90 per SD increase in the score for quantitative traits. We then assessed the adjusted predictive utility of the ITRS by fitting separate models for each phenotype and including its corresponding ITRS and PRS in the same model, adjusting for the same covariates (see **Methods**). Conditioning on PRS, all associations between ITRS-transcriptome and the phenotypes remained significant, with HRs ranging from 1.20 to 1.23 for disease phenotypes and betas ranging from 0.03 to 4.08 for quantitative traits. Similarly, all significant associations between ITRS-drug and the phenotypes remained, with HRs ranging from 1.07 to 1.09 for disease phenotypes and betas ranging from 0.01 to 1.50 for quantitative traits. In addition to effect size estimates, we obtained from each multivariate model the Harrell’s C-index^18^ (for disease phenotypes) or R-squared (for quantitative traits) as a measurement of model performance. The measurements were compared to baseline metrics obtained from multivariate models containing covariates only (**Figure 3, Supplemental Table 4**).

To show the discrimination of risk prediction scores, we classified each participant to different risk groups based on their score values. We defined three risk groups for each score, including a low (lowest 10% of the distribution), intermediate (middle 80% in the distribution), and high (highest 10% in the distribution) risk categories. Cumulative risk of disease phenotypes stratified by risk groups are presented in the format of Kaplan–Meier (KM) plots (**Figure 4** and **Supplemental Figure 2-3**) and distributions of quantitative traits across risk groups are presented in the format of box-plots (**Figure 5** and **Supplemental Figure 4-5**). We observed significant differences between risk groups and modest correlations between ITRS-transcriptome and ITRS-drug.

**Figure 4.**
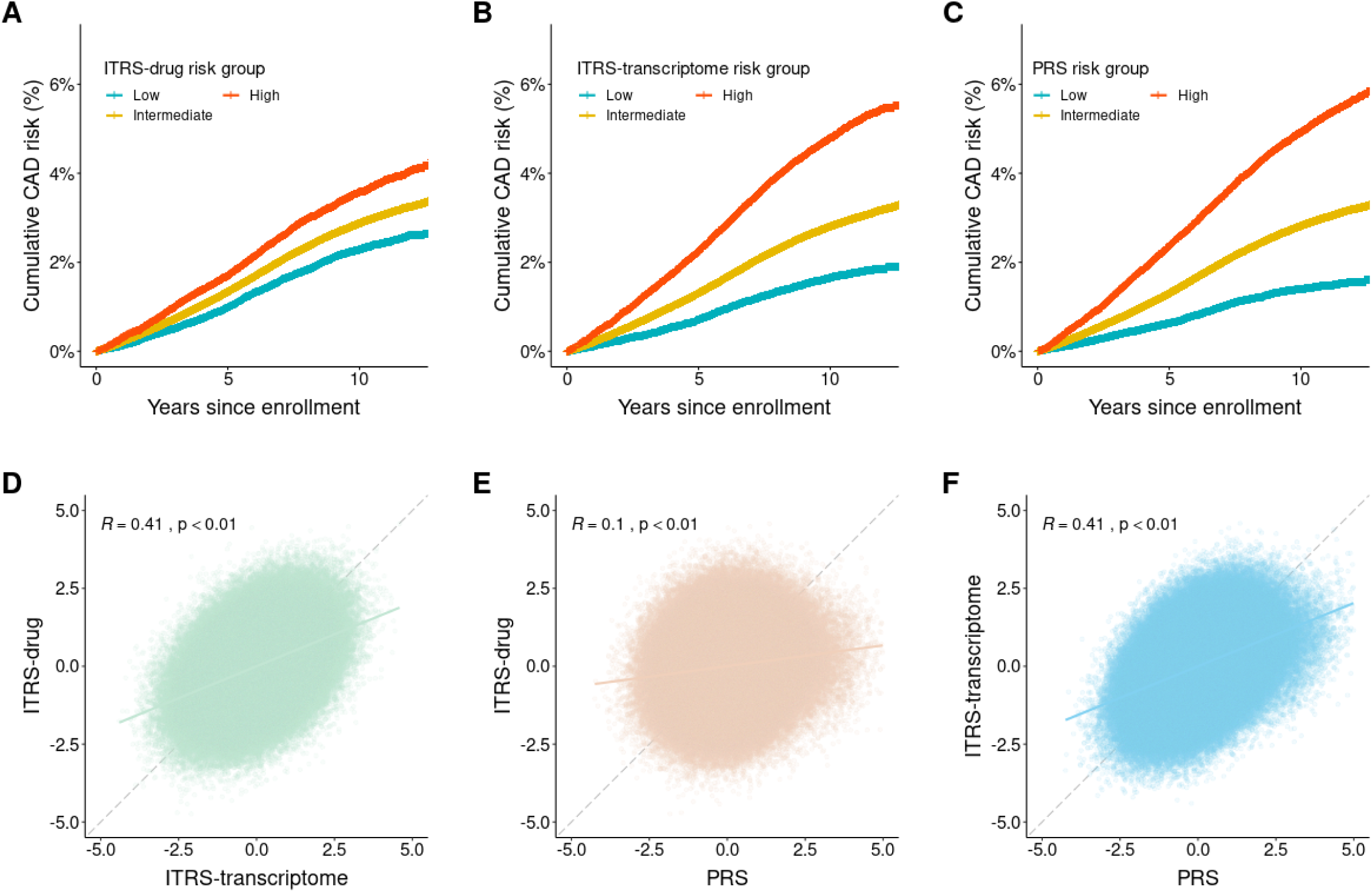
Risk prediction scores for coronary artery disease. **CAD:** coronary artery disease; **ITRS-transcriptome:**imputed transcriptome risk score constructed using suggestive significant tissue-gene pairs observed in transcriptome-wide association study (TWAS); **ITRS-drug:** ITRS constructed using a subset of suggestive significant tissue-gene pairs observed in TWAS, for which the genes are actionable proteins defined in a previous study;^14^ **PRS:** genome-wide polygenic risk score. All risk scores are plotted in standardized units calculated using the entire validation sample (N=471,600). **(A)**cumulative risk of CAD stratified by ITRS-drug risk groups (Low: lowest 10%, Intermediate: middle 80%, High: highest 10%); **(B)** cumulative risk of CAD stratified by ITRS-transcriptome risk groups; **(C)** cumulative risk of CAD stratified by PRS risk groups; **(D)** correlation between ITRS-transcriptome and ITRS-drug; ***R***: Pearson correlation coefficient; **(E)** correlation between PRS and ITRS-drug; **(F)**correlation between PRS and ITRS-transcriptome.

**Figure 5.**
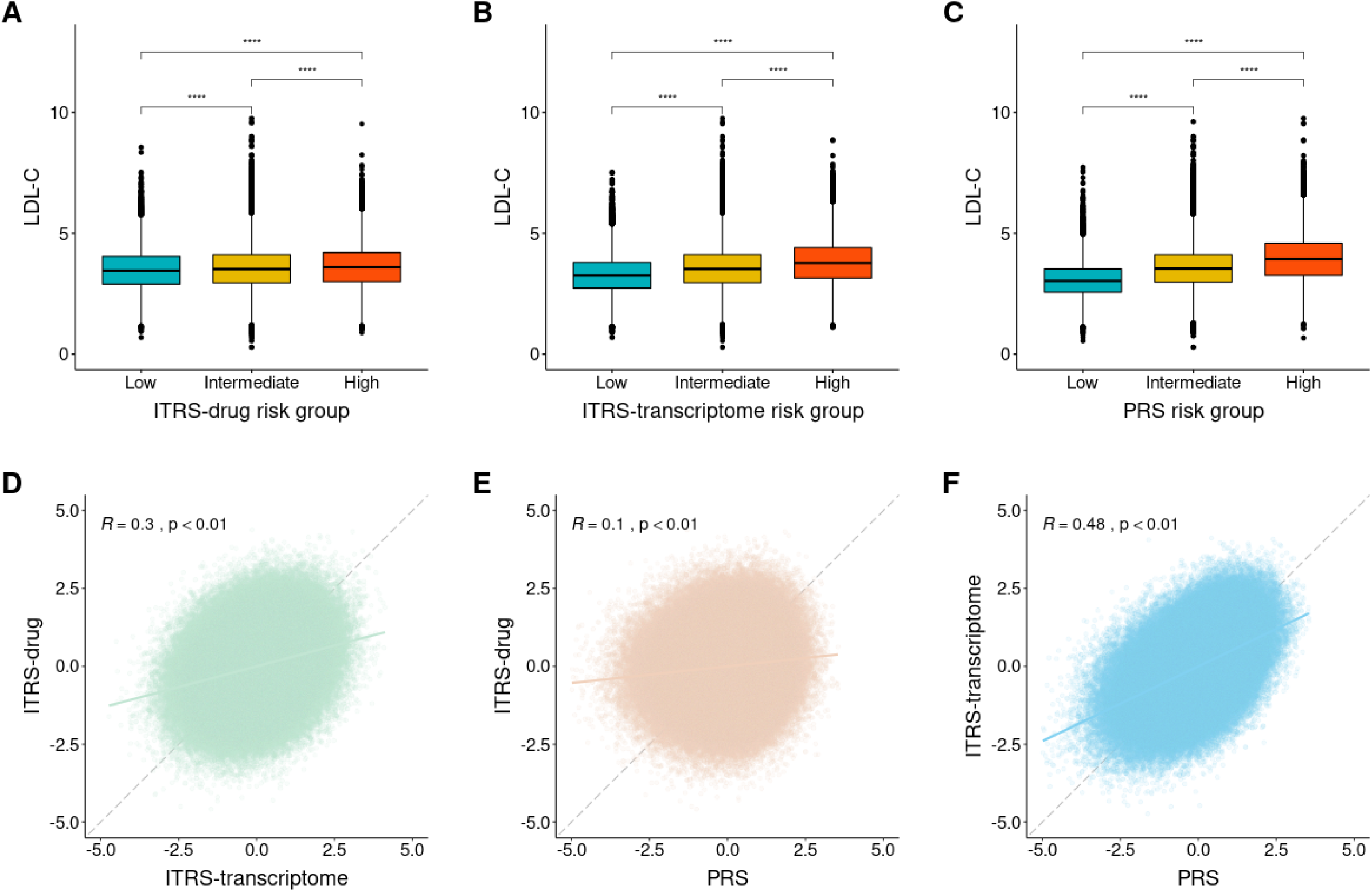
Risk prediction scores for low-density lipoprotein cholesterol. **LDL-C:** low-density lipoprotein cholesterol; **ITRS-transcriptome:** imputed transcriptome risk score constructed using suggestive significant tissue-gene pairs observed in transcriptome-wide association study (TWAS); **ITRS-drug:** ITRS constructed using a subset of suggestive significant tissue-gene pairs observed in TWAS, for which the genes are actionable proteins defined in a previous study;^14^ **PRS:** genome-wide polygenic risk score. All risk scores are plotted in standardized units calculated using the entire validation sample (N=323,931). **(A)** distributions of LDL-C levels across ITRS-drug risk groups (Low: lowest 10%, Intermediate: middle 80%, High: highest 10%); **(B)** distributions of LDL-C levels across ITRS-transcriptome risk groups; **(C)** distributions of LDL-C levels across PRS risk groups; **(D)** correlation between ITRS-transcriptome and ITRS-drug; ***R***: Pearson correlation coefficient; **(E)**correlation between PRS and ITRS-drug; **(F)** correlation between PRS and ITRS-transcriptome.

### PheWAS of established therapeutic targets using whole blood expression

In addition to the above phenotype-centered analysis, we conducted a gene-centered analysis to explore the potential of imputed gene expression for therapeutic targeting using high-throughput phenotyping. We focused the analysis on established gene targets of approved therapies, with the intent to both determine if on-target associations could be recapitulated, as well as if opportunities for repurposing of established drugs could be identified. After imputing expression levels of protein coding genes in whole blood (N=8,595), we identified 127 established therapeutic targets (genes encode a single protein target or known drug-binding subunit of a protein complex target) and retrieved indications of medications targeting these genes from the Open Targets Platform.^16^ We then manually mapped the retrieved indications to index phecodes to facilitate the downstream analysis in the framework of phecode-based PheWAS. In total, we identified 94 genes for which the indications can be mapped to at least one phecode, and associated their expression levels with 541 index phecodes representing largely independent phenotypes (**Supplemental Table 5**). As a generic tissue, whole blood was chosen to serve as a proxy for biologically relevant tissues corresponding to a variety of medications. The expression profile of whole blood was demonstrated to have moderate to high correlation with other tissues available in GTEx (average Pearson correlation coefficients: 0.66).^8^

In summary, 50,854 gene-phecode association tests were performed (94 genes tested for association with 541 phecodes). The potential on-target testing set comprised 324 gene-phecode pairs for which the phecode was consistent with an approved drug indication, and the potential repurposing testing set comprised 50,530 gene-phecode pairs for which the phecode did not reflect an approved drug indication. In the on-target test set, 3 (0.93%) of the 324 gene-phecode pairs were significant in the entire PheWAS at the FDR significance level of 0.05, and 30 (9.23%) had an unadjusted *P* value < 0.05 suggestive of association. In the potential repurposing test set, 12 of the 50,530 gene-phecode pairs (0.02%) were significant at the FDR significance level of 0.05, and 2,803 (5.55%) had an unadjusted *P* value < 0.05 suggestive of association. The 15 significant signals are shown in **Supplemental Figure 6**. Full results of the 50,854 gene-phecode association tests are provided in **Supplemental Table 6**. As with the TWAS approach above, we then performed locus-level colocalization analysis^11^ for significant associations, and observed 11 gene-phecode pairs had a non-zero colocalization probability, with 3 exceeding a colocalization probability > 0.50. Taking into account the mechanism of the existing therapeutic agents and the direction of lowering phenotypic risk, 6 gene-phecode pairs were considered to be good candidates, including 3 on-target replications and 3 potential repurposing opportunities (**Table 3**).

**Table 3.**
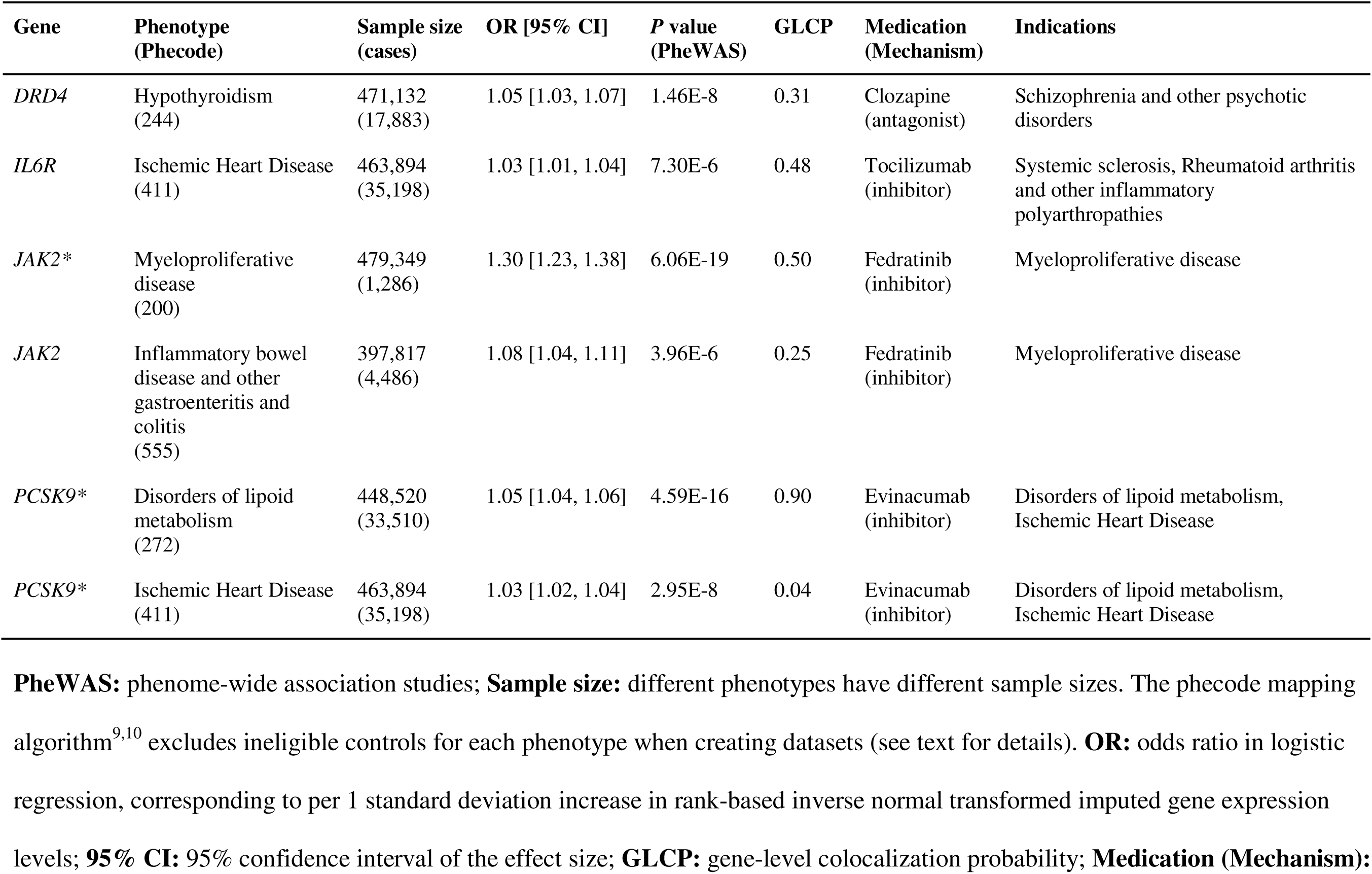
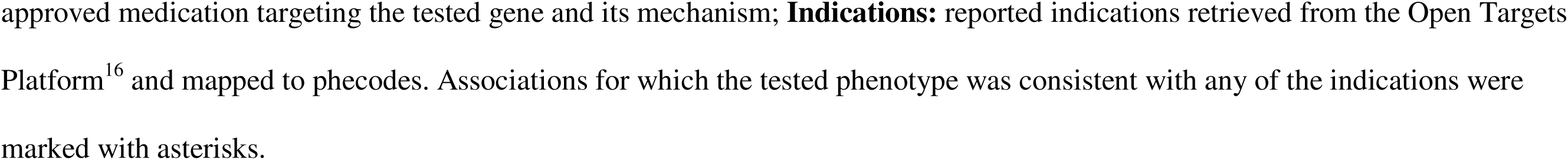
Whole blood PheWAS signals and colocalization probability.

| Gene | Phenotype<br>(Phecode) | Sample size<br>(cases) | OR [95% CI] | P value<br>(PheWAS) | GLCP | Medication<br>(Mechanism) | Indications |
| --- | --- | --- | --- | --- | --- | --- | --- |
| *DRD4* | Hypothyroidism<br>(244) | 471,132<br>(17,883) | 1.05 [1.03, 1.07] | 1.46E-8 | 0.31 | Clozapine<br>(antagonist) | Schizophrenia and other psychotic disorders |
| *IL6R* | Ischemic Heart Disease<br>(411) | 463,894<br>(35,198) | 1.03 [1.01, 1.04] | 7.30E-6 | 0.48 | Tocilizumab<br>(inhibitor) | Systemic sclerosis, Rheumatoid arthritis and other inflammatory polyarthropathies |
| *JAK2* * | Myeloproliferative disease<br>(200) | 479,349<br>(1,286) | 1.30 [1.23, 1.38] | 6.06E-19 | 0.50 | Fedratinib<br>(inhibitor) | Myeloproliferative disease |
| *JAK2* | Inflammatory bowel disease and other gastroenteritis and colitis<br>(555) | 397,817<br>(4,486) | 1.08 [1.04, 1.11] | 3.96E-6 | 0.25 | Fedratinib<br>(inhibitor) | Myeloproliferative disease |
| *PCSK9* * | Disorders of lipid metabolism<br>(272) | 448,520<br>(33,510) | 1.05 [1.04, 1.06] | 4.59E-16 | 0.90 | Evinacumab<br>(inhibitor) | Disorders of lipid metabolism, Ischemic Heart Disease |
| *PCSK9* * | Ischemic Heart Disease<br>(411) | 463,894<br>(35,198) | 1.03 [1.02, 1.04] | 2.95E-8 | 0.04 | Evinacumab<br>(inhibitor) | Disorders of lipid metabolism, Ischemic Heart Disease |

**PheWAS:** phenome-wide association studies; **Sample size:** different phenotypes have different sample sizes. The phecode mapping algorithm<sup>9,10</sup> excludes ineligible controls for each phenotype when creating datasets (see text for details). **OR:** odds ratio in logistic regression, corresponding to per 1 standard deviation increase in rank-based inverse normal transformed imputed gene expression levels; **95% CI:** 95% confidence interval of the effect size; **GLCP:** gene-level colocalization probability; **Medication (Mechanism):**
approved medication targeting the tested gene and its mechanism; **Indications:** reported indications retrieved from the Open Targets Platform<sup>16</sup> and mapped to phecodes. Associations for which the tested phenotype was consistent with any of the indications were marked with asterisks.

## Discussion

In the present study, we demonstrated specific attributes of individual-level imputed transcriptomes by showing three potential clinical and therapeutic applications in 486,452 UK Biobank participants. We imputed gene expression in six tissues that were biologically relevant to AF, CAD, T2D, QTc, LDL-C, or HbA1c. We then conducted tissue-specific TWAS for the phenotypes and identified 1,082 significant gene-tissue-phenotype trios after Bonferroni correction. 144 of the 1,082 signals were further supported by a colocalization analysis, among which 12 involved genes that were potential therapeutic targets. To evaluate the value of imputed transcriptomes in predicting phenotypic risk, we constructed two ITRS for each phenotype and observed consistent significant predictive abilities, even when conditioning on genome-wide PRS. Lastly, we performed a gene-centered analysis in which we ran PheWAS for 94 genes in whole blood that were targets of approved medications. We identified 6 good candidates with strong statistical evidence, including known indications and novel signals supported by previous literature and ongoing clinical trials.

These results have several implications. First, as a gene prioritization method focusing primarily on gene expression, the joint analysis of TWAS and colocalization can prioritize candidates for drug discovery or repurposing. Genes whose expression levels are significantly associated with phenotypic risk may indicate that the respective gene products or pathways in which they are involved serve as suitable targets for disease treatment. For example, one top signal in our joint analysis indicates that liver expression of *ANGPTL3* is positively associated with LDL-C. The encoded protein of *ANGPTL3* is predominantly expressed in the liver and is important for lipid metabolism. Indeed, *ANGPTL3* is the target of a group of new medications that inhibit its expression in the liver to treat hypercholesterolemia.^19^ Another interesting example is the positive association between atrial appendage *IL6R* expression and AF risk. *IL6R* encodes a subunit of the interleukin 6 receptor complex, and interleukin 6 is a key inflammatory cytokine. Inflammation has long been hypothesized to be a risk factor of AF,^20, 21^ and previous studies have associated SNPs in *IL6R* with AF risk.^22, 23^ Our results further support this association in heart tissues and imply that *IL6R* may be a potential therapeutic target for AF. We also noticed that the joint analysis missed some true biology when filtering out gene-tissue-phenotype trios with a low colocalization probability. For example, liver expression of *APOE* was significantly positively associated with LDL-C in TWAS, but the colocalization probability was 0.23. Similarly, left ventricle expression of *KCNQ1* was significantly negatively associated with QTc in TWAS, while the GLCP was 0.21. These false negatives may result from a failure to identify either the phenotype-SNP association or the expression-SNP association. Given the limited sample size in GTEx, the statistical power for identifying eQTL may be the issue. As sample sizes of gene expression panels increase, we anticipate improved power for eQTL and colocalization analysis and also better expression imputation in TWAS.

Second, ITRS can predict phenotypic risk in a biologically interpretable way and have significant predictive utility even when conditioning on PRS, perhaps identifying individuals most susceptible to disease via specific pathways. The predictors used in ITRS are expression levels of genes, which are more interpretable than genetic variants for which the mechanism underlying disease susceptibility may not be well understood. We observed consistent significant associations between the two ITRS and their corresponding phenotypes in the current study. When stratifying the study population into different risk groups based on their ITRS values, we observed significant separations of outcomes (cumulative risk for disease phenotypes, distributions for quantitative traits) between groups. We also observed that associations between ITRS and phenotypes remained significant and were largely unaffected by adjustment for a phenotype-specific genome-wide PRS, indicating that expression risk scores provide complementary predictive value to PRS.^24, 25^ Although genome-wide PRS outperformed ITRS in almost all scenarios we tested, it performed less well than ITRS-transcriptome in predicting QTc when using the same derivation and validation datasets. We hypothesize that risk prediction performance may differ according to the genetic architecture of specific phenotypes. Predicting phenotypic risk using genes that are targets of potential therapeutics (i.e., ITRS-drug) may help identify patients who are likely to benefit more from specific therapeutics. We observed only modest correlations between ITRS-transcriptome and ITRS-drug. Transcriptome-wide high-risk individuals who also scored high in ITRS-drug may benefit more from specific treatments targeting genes or pathways involved in the calculation of ITRS-drug.

Lastly, phenome-wide association analysis of potential therapeutically targetable genes may reveal novel associations that can guide indication expansion and facilitate prioritization of genes for drug development. In our PheWAS analysis focusing on 94 genes in whole blood deemed to be high-confidence therapeutic targets, 3 out of 324 (0.93%) associations testing a phenotype consistent with approved indications were significant and 12 out of 50,530 (0.02%) associations testing a phenotype inconsistent with approved indications were significant. Consistent associations indicate on-target replication and inconsistent associations prioritize expression-phenotype pairs for drug repurposing. For example, we observed that *JAK2* expression was significantly associated with myeloproliferative disease in PheWAS with a colocalization probability of 0.50, consistent with established indications. *JAK2* expression was also associated with inflammatory bowel disease, a potential novel indication. Although having a relatively low colocalization probability (GLCP: 0.25), an increased risk of inflammatory bowel disease has been reported in patients with myeloproliferative neoplasms^26^ and janus kinase antagonists have emerged as a new promising group of medications for treating inflammatory bowel disease.^27, 28^ We also noted that the significant association between *IL6R* and ischemic heart disease has a moderate colocalization probability of 0.48, further support the role of inflammation in cardiovascular disease.^29^

Our results should be interpreted in the context of design. First, the samples included in the current study were predominantly of European ancestry, which may limit the generalizability of our findings. Nevertheless, our observation of well-validated associations such as *PCSK9* and lipid disorders is reassuring, and we anticipate the robustness of association testing will improve as larger multi-ancestry samples become available. Second, the joint analysis of TWAS and colocalization can alternatively be conducted efficiently using summary statistics. However, individual-level TWAS is flexible to different covariate adjustments and gene expression prediction model assumptions, such as non-linearity of variant-expression associations. Third, we conducted the gene-centered PheWAS using imputed expression in whole blood only, which may result in false negative associations in cases in which gene expression-phenotype associations are tissue specific. We selected whole blood as a practical tissue to demonstrate feasibility of the approach, and submit that the approach is reasonable given the relative large sample for whole blood in GTEx and moderate to high correlation between whole blood expression with that of other tissues.^8^ We propose that future work examining precise tissue-level gene expression is warranted when considering drug repurposing or other therapeutic applications. Fourth, phenotype misclassification is possible given the fact that disease ascertainment relied on ICD-10 codes for the whole blood transcriptional PheWAS and that therapeutic indications in the Open Target Platform database may be imprecise or may not map directly to phecodes. For example, whereas the listed indications for *VKORC1* (the gene target for the anticoagulant warfarin) included atrial fibrillation, anticoagulants are used precisely for thromboembolism prevention, not treatment of atrial fibrillation itself. We anticipate that more precise phenotyping algorithms and drug indication resources will improve analytic resolution. Fifth, we acknowledge that gene expression may vary over time or in response to particular exposures, which are not routinely captured in tissue expression repositories and which may be prohibitive to model using longitudinal biobank data. Validation of associations with directly measured expression levels, when feasible, is warranted.

In conclusion, our results demonstrate potential clinical and therapeutic applications of individual-level imputed transcriptomes in a large, national, longitudinal biorepository. By conducting joint analysis of tissue-specific TWAS and locus-level colocalization, we recapitulate known biology and prioritize signals for drug repurposing opportunities. Using results from the TWAS, we constructed imputed transcriptome risk scores to predict phenotypic risk in a biologically interpretable manner. ITRS were shown to have significant predictive utility even when conditioning on genome-wide PRS. PheWAS of therapeutic targets using the rich phenotypic resources in large biorepositories like the UK biobank may enable prioritization of gene-phenotype associations to guide identification of novel disease indications.

## Online Methods

### Study sample

UK Biobank is a prospective cohort study with 502,629 participants aged 40-69 years at baseline recruited in the United Kingdom from 2006 to 2010.^30, 31^ Biological samples and detailed phenotypic data were collected for all participants who signed an electronic consent at enrollment. Specifically, the study collected a wide range of health-related variables, including survey data, physical measurements, longitudinal electronic health records, national registry, multimodal imaging, and laboratory test results. 488,377 participants underwent genome-wide genotyping as summarized below. Of these, 486,452 passed quality control (QC) procedures (reported gender consistent with genetically inferred gender; were not high genotype missingness rate or high heterozygosity rate outliers; were not sex chromosome aneuploidy) and have complete covariates data (enrollment age, sex, genotyping platform, and the top 5 principal components of ancestry) for further analysis. Use of UK Biobank data (under application number 17488) for the current study was approved by the Mass General Brigham (MGB) Institutional Review Board.

### Genomic and phenotypic data

Among the 488,377 genotyped participants, 49,950 were assayed at 807,411 genetic markers using the UK BiLEVE Axiom Array and 438,427 were assayed at 825,927 markers using the UK Biobank Axiom Array. The two genotyping arrays shared 95% marker content, which was chosen to capture single nucleotide polymorphism (SNPs) and short insertions and deletions (INDELs) at genome-wide scale.^31^ After variant-level and sample-level QC procedures, these variants were phased and imputed to the Haplotype Reference Consortium (HRC) and the merged UK 10K and 1000 Genomes phase III reference panels, producing nearly 96 million makers in the imputed genomic dataset.^31^ We used allele dosage from this imputed dataset to predict genetically regulated gene expression in the current study population.

We included three disease phenotypes (AF, CAD, T2D) and three quantitative traits (QTc, LDL-C, HbA1c) in the present study. For disease phenotypes, we have complete data for all 486,452 participants who passed QC procedures and were included in the imputation step. Combinations of international classification of diseases (ICD) 9 or 10 codes, procedure codes, and self-reported medical history were used to define disease phenotypes (**Supplemental Table 8**). Among the 486,452 individuals, 41,490, 462,739, and 462,159 had QTc, LDL-C, and HbA1c data available, respectively. QTc intervals were collected during the image visit when ECG tests were performed. LDL-C and HbA1c levels were measured at recruitment using blood biochemistry assays.

### Transcriptome imputation

We implemented a joint-tissue imputation (JTI) approach^8^ to predict the *cis* genetic component of gene expression for 486,452 UK Biobank participants in seven tissues, including heart atrial appendage, heart left ventricle, subcutaneous adipose, liver, coronary artery, pancreas, and whole blood. Inspired by the observation that gene expression profiles across different tissues are correlated and eQTL sharing is abundant,^3, 32^ JTI leverages the similarity of expression profiles and regulatory elements (calculated using the Encyclopedia of DNA Elements [ENCODE] project^33^ and Roadmap Epigenomics^34^ dataset) across tissues to improve transcriptome imputation in a target tissue. For each imputable gene in a given tissue, JTI uses a penalized regression model to select genetic variants (biallelic SNPs that passed quality control) within the *cis* window (window size varied by genes, determined based on cross-validation performance) that are significantly associated with expression levels in that tissue. The expression levels of each gene used in model derivation was the residuals of the normalized expression after adjustment (sex, sequencing platform, the top 5 principal components of ancestry, and Probabilistic Estimation of Expression Residuals [PEER] factors for each tissue). We obtained the pre-trained variant weights generated by these models from an open database [https://zenodo.org/record/3842289#.YnFfpvPMK3I] and computed the individual-level genetically regulated gene expression as a weighted allele score using PLINK 2.0 [https://www.coggenomics.org/plink/2.0/].

### Transcriptome-wide association studies and druggability definition

To perform individual-level transcriptome-wide association studies, we first applied rank-based inverse normal transformation (INT) to the predicted genetically regulated gene expression levels. We then associated each phenotype with the INT-transformed expression of each protein-coding gene in its biologically relevant tissues using regression models (logistic regression for binary traits, linear regression for quantitative traits), adjusting for enrollment age, sex, genotyping platform, and the top 5 principal components of ancestry. We determined significance using a Bonferroni corrected *P* value < 4.5×10^-7^ (0.05/110,109 tests) and suggestive significance using false discovery rate (adjusted *P* value < 0.01). All analyses were performed using R version 4.0. We adopted a dataset curated by Gaziano et al^14^ to determine the druggability of imputable protein-coding genes. The authors used the ChEMBL v.26 database^15^ to identify 1,263 actionable human proteins that were drug targets of approved or clinical-trial-stage medications. Indications for approved medications were retrieved from the Open Target Platform.^16^ Specifically, we downloaded the target-disease evidence database from the platform and restricted it to medications targeting the prioritized genes and having a ATC or FDA tag. Reported indications were then retrieved from the DailyMed [https://dailymed.nlm.nih.gov/dailymed/] entries corresponding to the identified medications.

### Locus-level colocalization analysis

We implemented a locus-level colocalization analysis approach proposed by Hukku et al^11^ to estimate the probability that causal variants for gene expression (eQTL) and the tested phenotype (hit SNPs) coexist in the same locus. The method differs from variant-level colocalization analysis in that it tests whether causal variants coexist in an intersected credible set generated by fine-mapping eQTL and hit SNPs, respectively. Members in the same credible set satisfy the redundancy and the LD conditions. Specifically, those variants, with only slight quantitative differences and in high LD, could represent the same underlying significant association nearly interchangeably. The development of this method was driven by the observed limited statistical power of variant-level colocalization analysis in practice.^11, 35, 36^

To estimate the gene-level colocalization probability for significant gene-tissue-phenotype trios identified in TWAS, we first subset the genetic data for each gene region of interest. Variants within 1 Mb upstream and 500 Kb downstream of the gene were extracted and estimated for associations with the tested phenotype using PLINK 2.0 [https://www.coggenomics.org/plink/2.0/]. The test statistics were fine-mapped to generate 95% credible sets using the FINEMAP software.^37^ LD information used in the fine-mapping procedure was in-sample LD calculated from the subsetted genetic data using the LDstore software.^38^ The fine-mapped results were then integrated with pre-computed GTEx multi-tissue eQTL annotation to estimate the locus-level colocalization probability in the tested tissue using the fastENLOC software.^11^ eQTL annotations were generated from software package DAP-G.^39^

### Construction and validation of imputed transcriptome risk scores

We constructed two imputed transcriptome risk scores (ITRS) for each phenotype, one with all imputable protein-coding genes identified at suggestive significance level (adjusted *P* value from association test < 0.01), another with a subset of druggable genes identified at the same level. For each disease phenotype, we first obtained scoring weights using a prevalent dataset in the UK Biobank, in which participants who were diagnosed with the disease before enrollment were defined as cases. All participants who were free of disease at baseline were identified as referents, including those who developed the disease during follow-up. Using the derivation dataset, we input (1) all suggestive significant gene-tissue pairs (e.g., *TBX5* expression in the left ventricle for AF) and (2) a subset of (1), consisting of potential druggable genes, into two separate elastic net regression models (□ = 0.5, □ was determined through 10 folds cross-validation) to adjust scoring weights and address potential correlations between predictors. R package *glmnet* was used to fit the models. We transformed the imputed expression values using rank-based inverse normal transformation and regressed covariates (enrollment age, sex, genotyping platform, and top 5 principal components of ancestry) out before modeling and scoring. After obtaining the weights from penalized regression models, we calculated and validated the ITRS for each disease phenotype using an incident dataset in the UK Biobank, in which all prevalent cases were removed. The association between each disease phenotype and its corresponding ITRS (standardized) was evaluated using a Cox proportional-hazards regression model, adjusting for enrollment age, sex, genotyping platform, and top 5 principal components of ancestry. Time to disease diagnosis was defined using the longitudinal information available in the UK Biobank, with censoring at last follow-up. This analysis framework has been described and implemented in previous studies.^40, 41^ The two steps consider different periods of risk and thus can be seen as relatively independent.

We used a similar strategy to construct and validate ITRS for quantitative traits, except that the derivation and validation datasets were different from what we created for disease phenotypes. For each quantitative trait, we first randomly split the complete dataset into a derivation set (30%) and a validation set (70%). We then obtained scoring weights using an elastic net model in the derivation set and calculated and validated the ITRS in the validation set using a linear regression model, adjusting for the same covariates.

### Calculation and testing of polygenic risk scores

To make standardized comparisons, instead of using external large-scale GWAS summary statistics to create polygenic risk scores (PRS) for the phenotypes, we ran GWAS in the derivation sets we used for ITRS to obtain allele effect estimates for PRS. We used a whole-genome regression approach implemented in the REGENIE^42^ v2.0.2 software to test the association between each phenotype and individual SNPs assuming an additive genetic model, adjusting for enrollment age, sex, genotyping platform, and top 5 principal components of ancestry. For binary traits, we additionally accounted for case-control imbalance using the saddle point approximation (SPA) method^43^ implemented in REGENIE. We then obtained scoring weights and calculated PRS in the same ITRS validation datasets using PRS-CS-auto.^17^ This method implements a fully Bayesian framework, which includes in score calculation all SNPs in the GWAS summary statistics and applies shrinkage factors to their original allele effect estimates to avoid overfitting. The PRS were calculated as the PRS-CS-auto adjusted SNP effect estimate-weighted sum of genetic dosage at effect alleles using PLINK 2.0 [https://www.coggenomics.org/plink/2.0/].

We tested the utility of PRS similarly to ITRS. Specifically, we evaluated the association between each phenotype and its corresponding PRS (standardized) in the validation dataset using regression models (Cox models for disease phenotypes, linear regression for quantitative traits), adjusting for the same covariates. To evaluate the independent predictive value of ITRS, in addition to the regression models described above, we fitted separate models for each phenotype and included its corresponding ITRS and PRS in the same model to assess their relative contribution as predictors, adjusting for the same covariates.

### Measurement of model performance

We used Harrell’s C-index as the model performance metric for disease phenotypes. Harrell’s C-index is a concordance index commonly used in survival analysis to measure the performance of risk scores in discriminating patients with different risks.^18^ We first calculated a C-index in a baseline model containing covariates only (enrollment age, sex, genotyping platform, and the top 5 principal components of ancestry), we then compared it to C statistics obtained from multivariate Cox models involving risk scores as described above. For quantitative traits, we used R-squared as the model performance metric. R-squared measures the variation in the quantitative trait explained by modeling variables. Similarly, we fitted a linear regression model with covariates only to obtain a baseline estimate and compared it to the R-squared obtained from multivariate linear regression models involving risk scores as described above. R packages *survival* and *stats* were used to fit the models and obtain the performance metrics.

### PheWAS established therapeutic targets in whole blood

In addition to the phenotype-centered analysis described above, we included in our study a gene-centered analysis to demonstrate a potential use case of individual-level imputed transcriptomes that may be difficult to implement using GWAS summary statistics. Using the actionable protein data cited above,^14^ we selected a subset of genes that were established targets of approved medications and imputed their *cis* genetic component of gene expression in whole blood. We then ran a phenome-wide association study (PheWAS) for each imputed druggable gene in whole blood to search for novel associations implicating drug repurposing opportunities. As a generic tissue, whole blood was chosen to serve as a proxy for biological relevant tissues corresponding to a variety of medications included in the analysis.

To create phenotypes for the PheWAS analysis, we used the Phecode mapping algorithm^9, 10^ implemented in the R package *PheWAS* with the ICD-10 data in the UK Biobank and generated 1,590 disease phenotypes with non-zero cases (defined as having at least one eligible ICD code). The algorithm groups ICD codes representing the same disease together (e.g., primary and secondary codes) to improve the statistical power of PheWAS and reduce the number of tests included in analysis. Motivated by the observation that some of the resulting phecodes were highly correlated (e.g., code 272.10 represents hyperlipidemia and code 272.11 represents hypercholesterolemia) and therefore may result in redundant tests, we applied a hierarchical clustering algorithm to the 1,590 phenotypes and identified a subset of 629 largely independent phenotypes for downstream analysis. Specifically, we first constructed a cosine similarity matrix and a cosine distance matrix (1-similarity matrix) using all phecodes. We then applied Ward’s method^44^ to the distance matrix for hierarchical clustering. Phecode clusters were defined using a clustering tree height cut-off of 1.0, and index phecode was identified as the phenotype with the highest number of cases within each cluster. For each imputed gene (rank-based inverse normal transformed), we tested the associations between imputed expression levels and 541 major phenotypes (>20 cases) using logistic regression, adjusting for age, sex, genotyping platform, and the top 5 principal components of ancestry. We used false discovery rate (adjusted *P* value < 0.05) to determine the significance of associations and for each significant gene we retrieved the indications of approved medications targeting it from the Open Target Platform.^16^ The indications were then manually mapped to the index phecodes to facilitate downstream analysis in the framework of Phecode-PheWAS.

## Data Availability

Access to individual-level genetic and phenotypic data in the UK Biobank (UKB) is available to researchers through application on the UKB website (https://www.ukbiobank.ac.uk). Use of UKB data in the current study was performed under application number 17488.

## References

1. Li B, Ritchie MD. From GWAS to Gene: Transcriptome-Wide Association Studies and Other Methods to Functionally Understand GWAS Discoveries. Front Genet. 2021;12. Accessed April 21, 2022. https://www.frontiersin.org/article/10.3389/fgene.2021.713230

2. Wainberg M, Sinnott-Armstrong N, Mancuso N, et al. Opportunities and challenges for transcriptome-wide association studies. Nat Genet. 2019;51(4):592–599. doi:10.1038/s41588-019-0385-z

3. THE GTEX CONSORTIUM. The GTEx Consortium atlas of genetic regulatory effects across human tissues. Science. 2020;369(6509):1318–1330. doi:10.1126/science.aaz1776

4. Gamazon ER, Wheeler HE, Shah KP, et al. A gene-based association method for mapping traits using reference transcriptome data. Nat Genet. 2015;47(9):1091–1098. doi:10.1038/ng.3367

5. Gusev A, Ko A, Shi H, et al. Integrative approaches for large-scale transcriptome-wide association studies. Nat Genet. 2016;48(3):245–252. doi:10.1038/ng.3506

6. Gusev A, Mancuso N, Won H, et al. Transcriptome-wide association study of schizophrenia and chromatin activity yields mechanistic disease insights. Nat Genet. 2018;50(4):538–548. doi:10.1038/s41588-018-0092-1

7. Hu Y, Li M, Lu Q, et al. A statistical framework for cross-tissue transcriptome-wide association analysis. Nat Genet. 2019;51(3):568–576. doi:10.1038/s41588-019-0345-7

8. Zhou D, Jiang Y, Zhong X, Cox NJ, Liu C, Gamazon ER. A unified framework for joint-tissue transcriptome-wide association and Mendelian randomization analysis. Nat Genet. 2020;52(11):1239–1246. doi:10.1038/s41588-020-0706-2

9. Denny JC, Bastarache L, Ritchie MD, et al. Systematic comparison of phenome-wide association study of electronic medical record data and genome-wide association study data. Nat Biotechnol. 2013;31(12):1102–1111. doi:10.1038/nbt.2749

10. Wu P, Gifford A, Meng X, et al. Mapping ICD-10 and ICD-10-CM Codes to Phecodes: Workflow Development and Initial Evaluation. JMIR Med Inform. 2019;7(4):e14325. doi:10.2196/14325

11. Hukku A, Sampson MG, Luca F, Pique-Regi R, Wen X. Analyzing and reconciling colocalization and transcriptome-wide association studies from the perspective of inferential reproducibility. Am J Hum Genet. 2022;109(5):825–837. doi:10.1016/j.ajhg.2022.04.005

12. Harrow J, Frankish A, Gonzalez JM, et al. GENCODE: the reference human genome annotation for The ENCODE Project. Genome Res. 2012;22(9):1760–1774. doi:10.1101/gr.135350.111

13. Mancuso N, Freund MK, Johnson R, et al. Probabilistic fine-mapping of transcriptome-wide association studies. Nat Genet. 2019;51(4):675–682. doi:10.1038/s41588-019-0367-1

14. Gaziano L, Giambartolomei C, Pereira AC, et al. Actionable druggable genome-wide Mendelian randomization identifies repurposing opportunities for COVID-19. Nat Med. 2021;27(4):668–676. doi:10.1038/s41591-021-01310-z

15. Gaulton A, Bellis LJ, Bento AP, et al. ChEMBL: a large-scale bioactivity database for drug discovery. Nucleic Acids Res. 2012;40(Database issue):D1100–D1107. doi:10.1093/nar/gkr777

16. Ochoa D, Hercules A, Carmona M, et al. Open Targets Platform: supporting systematic drug–target identification and prioritisation. Nucleic Acids Res. 2021;49(D1):D1302–D1310. doi:10.1093/nar/gkaa1027

17. Ge T, Chen CY, Ni Y, Feng YCA, Smoller JW. Polygenic prediction via Bayesian regression and continuous shrinkage priors. Nat Commun. 2019;10(1):1776. doi:10.1038/s41467-019-09718-5

18. Evaluating the Yield of Medical Tests | JAMA | JAMA Network. Accessed August 2, 2022. https://jamanetwork.com/journals/jama/article-abstract/372568

19. Reeskamp LF, Millar JS, Wu L, et al. ANGPTL3 Inhibition With Evinacumab Results in Faster Clearance of IDL and LDL apoB in Patients With Homozygous Familial Hypercholesterolemia—Brief Report. Arterioscler Thromb Vasc Biol. 2021;41(5):1753–1759. doi:10.1161/ATVBAHA.120.315204

20. Aviles RJ, Martin DO, Apperson-Hansen C, et al. Inflammation as a risk factor for atrial fibrillation. Circulation. 2003;108(24):3006–3010. doi:10.1161/01.CIR.0000103131.70301.4F

21. Issac TT, Dokainish H, Lakkis NM. Role of inflammation in initiation and perpetuation of atrial fibrillation: a systematic review of the published data. J Am Coll Cardiol. 2007;50(21):2021–2028. doi:10.1016/j.jacc.2007.06.054

22. Schnabel RB, Kerr KF, Lubitz SA, et al. Large-Scale Candidate Gene Analysis in Whites and African Americans Identifies IL6R Polymorphism in Relation to Atrial Fibrillation. Circ Cardiovasc Genet. 2011;4(5):557–564. doi:10.1161/CIRCGENETICS.110.959197

23. Cupido AJ, Asselbergs FW, Natarajan P, et al. Dissecting the IL-6 pathway in cardiometabolic disease: A Mendelian randomization study on both IL6 and IL6R. Br J Clin Pharmacol. 2022;88(6):2875–2884. doi:10.1111/bcp.15191

24. Liang Y, Pividori M, Manichaikul A, et al. Polygenic transcriptome risk scores (PTRS) can improve portability of polygenic risk scores across ancestries. Genome Biol. 2022;23(1):23. doi:10.1186/s13059-021-02591-w

25. Pain O, Glanville KP, Hagenaars S, et al. Imputed gene expression risk scores: a functionally informed component of polygenic risk. Hum Mol Genet. 2021;30(8):727–738. doi:10.1093/hmg/ddab053

26. Bak M, Jess T, Flachs EM, Zwisler AD, Juel K, Frederiksen H. Risk of Inflammatory Bowel Disease in Patients with Chronic Myeloproliferative Neoplasms: A Danish Nationwide Cohort Study. Cancers. 2020;12(9):2700. doi:10.3390/cancers12092700

27. Boland BS, Sandborn WJ, Chang JT. Update on Janus Kinase Antagonists in Inflammatory Bowel Disease. Gastroenterol Clin North Am. 2014;43(3):603–617. doi:10.1016/j.gtc.2014.05.011

28. Rogler G. Efficacy of JAK inhibitors in Crohn’s Disease. J Crohns Colitis. 2020;14(Supplement_2):S746–S754. doi:10.1093/ecco-jcc/jjz186

29. Ferrucci L, Fabbri E. Inflammageing: chronic inflammation in ageing, cardiovascular disease, and frailty. Nat Rev Cardiol. 2018;15(9):505–522. doi:10.1038/s41569-018-0064-2

30. Sudlow C, Gallacher J, Allen N, et al. UK Biobank: An Open Access Resource for Identifying the Causes of a Wide Range of Complex Diseases of Middle and Old Age. PLOS Med. 2015;12(3):e1001779. doi:10.1371/journal.pmed.1001779

31. Bycroft C, Freeman C, Petkova D, et al. The UK Biobank resource with deep phenotyping and genomic data. Nature. 2018;562(7726):203–209. doi:10.1038/s41586-018-0579-z

32. Aguet F, Brown AA, Castel SE, et al. Genetic effects on gene expression across human tissues. Nature. 2017;550(7675):204–213. doi:10.1038/nature24277

33. Davis CA, Hitz BC, Sloan CA, et al. The Encyclopedia of DNA elements (ENCODE): data portal update. Nucleic Acids Res. 2018;46(D1):D794–D801. doi:10.1093/nar/gkx1081

34. Kundaje A, Meuleman W, Ernst J, et al. Integrative analysis of 111 reference human epigenomes. Nature. 2015;518(7539):317–330. doi:10.1038/nature14248

35. Hukku A, Pividori M, Luca F, Pique-Regi R, Im HK, Wen X. Probabilistic colocalization of genetic variants from complex and molecular traits: promise and limitations. Am J Hum Genet. 2021;108(1):25–35. doi:10.1016/j.ajhg.2020.11.012

36. Schaid DJ, Chen W, Larson NB. From genome-wide associations to candidate causal variants by statistical fine-mapping. Nat Rev Genet. 2018;19(8):491–504. doi:10.1038/s41576-018-0016-z

37. Benner C, Spencer CCA, Havulinna AS, Salomaa V, Ripatti S, Pirinen M. FINEMAP: efficient variable selection using summary data from genome-wide association studies. Bioinformatics. 2016;32(10):1493–1501. doi:10.1093/bioinformatics/btw018

38. Benner C, Havulinna AS, Järvelin MR, Salomaa V, Ripatti S, Pirinen M. Prospects of Fine-Mapping Trait-Associated Genomic Regions by Using Summary Statistics from Genome-wide Association Studies. Am J Hum Genet. 2017;101(4):539–551. doi:10.1016/j.ajhg.2017.08.012

39. Wen X, Lee Y, Luca F, Pique-Regi R. Efficient Integrative Multi-SNP Association Analysis via Deterministic Approximation of Posteriors. Am J Hum Genet. 2016;98(6):1114–1129. doi:10.1016/j.ajhg.2016.03.029

40. Benjamin EJ, Rice KM, Arking DE, et al. Variants in ZFHX3 are associated with atrial fibrillation in individuals of European ancestry. Nat Genet. 2009;41(8):879–881. doi:10.1038/ng.416

41. Christophersen IE, Rienstra M, Roselli C, et al. Large-scale analyses of common and rare variants identify 12 new loci associated with atrial fibrillation. Nat Genet. 2017;49(6):946–952. doi:10.1038/ng.3843

42. Mbatchou J, Barnard L, Backman J, et al. Computationally efficient whole-genome regression for quantitative and binary traits. Nat Genet. 2021;53(7):1097–1103. doi:10.1038/s41588-021-00870-7

43. Zhou W, Nielsen JB, Fritsche LG, et al. Efficiently controlling for case-control imbalance and sample relatedness in large-scale genetic association studies. Nat Genet. 2018;50(9):1335–1341. doi:10.1038/s41588-018-0184-y

44. Ward JH. Hierarchical Grouping to Optimize an Objective Function. J Am Stat Assoc. 1963;58(301):236–244. doi:10.1080/01621459.1963.10500845

