## Supplementary Materials for "Clinical and Therapeutic Applications of Individual-level Tissue-Specific Imputed Transcriptomes"

#### Supplemental Materials

**Supplemental Table 4. Predictive utility of imputed transcriptome risk scores (ITRS) and polygenic risk scores (PRS) for select phenotypes** **2**

**Supplemental Table 8. Phenotype definitions of curated disease phenotypes** **6**

**Supplemental Figure 1. Training-testing framework for Imputed Transcriptome Risk Scores** **8**

**Supplemental Figure 2. Risk prediction scores for atrial fibrillation** **9**

**Supplemental Figure 3. Risk prediction scores for type 2 diabetes** **11**

**Supplemental Figure 4. Risk prediction scores for QTc interval on ECGs** **13**

**Supplemental Figure 5. Risk prediction scores for glycated hemoglobin** **15**

**Supplemental Figure 6. Genes representing therapeutic targets with at least one significant association identified in the whole blood phenome-wide association study** **17**

### Supplemental Table 4. Predictive utility of imputed transcriptome risk scores (ITRS) and polygenic risk scores (PRS) for select phenotypes

| | **Single Score Models** | | **ITRS-transcriptome and PRS Adjusted Models** | | **ITRS-drug and PRS Adjusted Models** | |
|---|---|---|---|---|---|---|
| **Diseases** | | | | | | |
| | **HR [95% CI] (Harrell's C-index)** | **Z score (*P* value)** | **HR [95% CI] (Harrell's C-index)** | **Z score (*P* value)** | **HR [95% CI] (Harrell's C-index)** | **Z score (*P* value)** |
| **AF (N=478,165 in test set)** | | | | | | |
| ITRS-transcriptome (197 genes) | 1.30 [1.28, 1.32] (0.75/0.74) | 39.76 (*P* < 1.00E-308) | 1.20 [1.18, 1.22] (0.76/0.74) | 26.65 (*P* = 1.99E-156) | | |
| ITRS-drug (13 genes) | 1.09 [1.08, 1.11] (0.74/0.74) | 13.41 (*P* = 5.64E-41) | | | 1.08 [1.06, 1.09] (0.75/0.74) | 11.18 (*P* = 5.18E-29) |
| PRS (genome-wide) | 1.39 [1.37, 1.41] (0.75/0.74) | 52.35 (*P* < 1.00E-308) | 1.33 [1.31, 1.34] (0.76/0.74) | 42.71 (*P* < 1.00E-308) | 1.39 [1.37, 1.40] (0.75/0.74) | 51.79 (*P* < 1.00E-308) |
| **CAD (N=471,600 in test set)** | | | | | | |
| ITRS-transcriptome (254 genes) | 1.39 [1.36, 1.41] (0.74/0.72) | 39.34 (*P* < 1.00E-308) | 1.23 [1.21, 1.25] (0.75/0.72) | 22.90 (*P* = 4.32E-116) | | |
| ITRS-drug (37 genes) | 1.15 [1.13, 1.17] (0.73/0.72) | 16.59 (*P* = 8.69E-62) | | | 1.09 [1.07, 1.11] (0.75/0.72) | 10.49 (*P* = 9.61E-26) |
| PRS (genome-wide) | 1.45 [1.43, 1.47] (0.74/0.72) | 47.06 (*P* < 1.00E-308) | 1.34 [1.32, 1.36] (0.75/0.72) | 33.52 (*P* = 2.70E-246) | 1.43 [1.41, 1.46] (0.75/0.72) | 45.12 (*P* < 1.00E-308) |

| | | | | | | |
|---|---|---|---|---|---|---|
| **T2D (N=474,481 in test set)** | | | | | | |
| ITRS-transcriptome (325 genes) | 1.34 [1.32, 1.36] (0.69/0.67) | 45.37 (*P* < 1.00E-308) | 1.21 [1.20, 1.23] (0.71/0.67) | 28.70 (*P* = 4.25E-181) | | |
| ITRS-drug (35 genes) | 1.13 [1.11, 1.14] (0.67/0.67) | 18.42 (*P* = 8.92E-76) | | | 1.07 [1.06, 1.08] (0.71/0.67) | 10.28 (*P* = 9.09E-25) |
| PRS (genome-wide) | 1.51 [1.49, 1.53] (0.70/0.67) | 64.65 (*P* < 1.00E-308) | 1.43 [1.41, 1.45] (0.71/0.67) | 54.15 (*P* < 1.00E-308) | 1.49 [1.48, 1.51] (0.71/0.67) | 62.80 (*P* < 1.00E-308) |
| **Quantitative Traits** | | | | | | |
| | **Beta [95% CI] (R squared)** | **T value (*P* value)** | **Beta [95% CI] (R squared)** | **T value (*P* value)** | **Beta [95% CI] (R squared)** | **T value (*P* value)** |
| **LDL-C (N=323,931 in test set)** | | | | | | |
| ITRS-transcriptome (308 genes) | 0.15 [0.14, 0.15] (0.04/0.01) | 97.99 (*P* < 1.00E-308) | 0.03 [0.03, 0.04] (0.09/0.01) | 19.00 (*P* = 1.84E-80) | | |
| ITRS-drug (22 genes) | 0.04 [0.04, 0.04] (0.01/0.01) | 27.22 (*P* = 6.14E-163) | | | 0.01 [0.01, 0.02] (0.09/0.01) | 9.56 (*P* = 1.17E-21) |
| PRS (genome-wide) | 0.26 [0.25, 0.26] (0.09/0.01) | 171.05 (*P* < 1.00E-308) | 0.24 [0.24, 0.24] (0.09/0.01) | 139.55 (*P* < 1.00E-308) | 0.25 [0.25, 0.26] (0.09/0.01) | 168.97 (*P* < 1.00E-308) |
| **HbA1c (N=323,542 in test set)** | | | | | | |
| ITRS-transcriptome (799 genes) | 0.99 [0.97, 1.02] (0.08/0.06) | 86.99 (*P* < 1.00E-308) | 0.52 [0.49, 0.54] (0.11/0.06) | 41.72 (*P* < 1.00E-308) | | |
| ITRS-drug (93 genes) | 0.36 [0.34, 0.39] (0.06/0.06) | 31.55 (*P* = 3.30E-218) | | | 0.12 [0.10, 0.15] (0.10/0.06) | 10.88 (*P* = 1.51E-27) |
| PRS (genome-wide) | 1.40 [1.38, 1.43] (0.10/0.06) | 120.78 (*P* < 1.00E-308) | 1.18 [1.16, 1.21] (0.11/0.06) | 92.94 (*P* < 1.00E-308) | 1.38 [1.36, 1.40] (0.10/0.06) | 116.94 (*P* < 1.00E-308) |
| **QTc (N=29,053 in test set)** | | | | | | |

| | | | | | | |
|---|---|---|---|---|---|---|
| ITRS-transcriptome (55 genes) | 4.98 [4.69, 5.26] (0.10/0.06) | 34.69 (*P* = 2.01E-258) | 4.08 [3.75, 4.41] (0.10/0.06) | 24.46 (*P* = 8.65E-131) | | |
| ITRS-drug (6 genes) | 2.39 [2.11, 2.68] (0.07/0.06) | 16.42 (*P* = 2.76E-60) | | | 1.50 [1.21, 1.79] (0.08/0.06) | 10.02 (*P* = 1.32E-23) |
| PRS (genome-wide) | 3.90 [3.62, 4.19] (0.08/0.06) | 26.55 (*P* = 1.85E-153) | 1.78 [1.44, 2.11] (0.10/0.06) | 10.46 (*P* = 1.40E-25) | 3.51 [3.21, 3.81] (0.08/0.06) | 23.09 (*P* = 6.31E-117) |

**ITRS-transcriptome:** imputed transcriptome risk score constructed using suggestive significant tissue-gene pairs observed in transcriptome-wide association study (TWAS); **ITRS-drug:** ITRS constructed using a subset of suggestive significant tissue-gene pairs observed in TWAS, for which the genes are actionable proteins defined in a previous study; **Single Score Models:** risk score was tested individually in a model for association with the corresponding phenotype, adjusting for age, sex, genotyping platform, and the top 5 principal components of ancestry. **ITRS-transcriptome and PRS Adjusted Models:** IRTS-all and PRS were tested together in a model, adjusting for the same covariates; **ITRS-drug and PRS Adjusted Models:** ITRS-drug and PRS were tested together in a model, adjusting for the same covariates; **HR:** hazard ratio in Cox proportional hazard models, corresponding to 1 standard deviation (SD) increase in risk scores; **Harrell's C-index:** concordance index obtained from the same model (estimate before slash), comparing to the baseline C-index obtained from a model containing only covariates (estimate after slash); **Beta:** effect size estimates in linear regression models, corresponding to 1 SD increase in risk scores; **R squared**: variation in traits explained by variables obtained from the same model (estimate before slash), comparing to the baseline R squared obtained from only covariates model (estimate after slash); **95% CI:** 95% confidence intervals; **AF:** atrial fibrillation; **CAD:** coronary artery disease; **T2D:** type 2 diabetes; **QTc:** heart rate-corrected QT interval on electrocardiograms; **LDL-C:** low-density lipoprotein cholesterol; **HbA1c:** hemoglobin A1C (glycated hemoglobin).

### Supplemental Table 8. Phenotype definitions of curated disease phenotypes

| Phenotype | Case definition | |
|---|---|---|
| | **UKB data field** | **Coding** |
| Atrial fibrillation | 20002 (self-report) | 1471,1483 |
| | 20004 (Operation code) | 1524 |
| | 41202 (ICD-10) | I48,I48.0,I48.1,I48.2,I48.3,I48.4,I48.9 |
| | 41204 (ICD-10) | I48,I48.0,I48.1,I48.2,I48.3,I48.4,I48.9 |
| | 40001 (ICD-10) | I48,I48.0,I48.1,I48.2,I48.3,I48.4,I48.9 |
| | 40002 (ICD-10) | I48,I48.0,I48.1,I48.2,I48.3,I48.4,I48.9 |
| | 41203 (ICD-9) | 4273 |
| | 41205 (ICD-9)) | 4273 |
| | 41200 (OPCS4) | K57.1,K62.1,K62.2,K62.3,K62.4,X50.1,X50.2 |
| | 41210 (OPCS4) | K57.1,K62.1,K62.2,K62.3,K62.4,X50.1,X50.2 |
| Coronary artery disease | 41200 (OPCS4) | K40.1,K40.2,K40.3,K40.4,K41.1,K41.2,K41.3,K41.4,K45.1,K45.2,K45.3,K45.4,K45.5,K49.1,K49.2,K49.8,K49.9,K50.2,K75.1,K75.2,K75.3,K75.4,K75.8,K75.9 |
| | 41210 (OPCS4) | K40.1,K40.2,K40.3,K40.4,K41.1,K41.2,K41.3,K41.4,K45.1,K45.2,K45.3,K45.4,K45.5, |

| | | |
|---|---|---|
| | 42001 (Source of myocardial infarction report) | K49.1,K49.2,K49.8,K49.9,K50.2,K75.1,K75.2,K75.3,K75.4,K75.8,K75.9<br><br>0,1,2 |
| Type 2 diabetes | 20002 (self-report)<br><br>41202 (ICD-10)<br><br>41204 (ICD-10)<br><br>40001 (ICD-10)<br><br>40002(ICD-10) | 1223<br><br>E11,E11.0,E11.1,E11.2,E11.3,E11.4,E11.5,E11.6,E11.7,E11.8,E11.9<br><br>E11,E11.0,E11.1,E11.2,E11.3,E11.4,E11.5,E11.6,E11.7,E11.8,E11.9<br><br>E11,E11.0,E11.1,E11.2,E11.3,E11.4,E11.5,E11.6,E11.7,E11.8,E11.9<br><br>E11,E11.0,E11.1,E11.2,E11.3,E11.4,E11.5,E11.6,E11.7,E11.8,E11.9 |

Supplemental Figure 1. Derivation-validation framework for Imputed Transcriptome Risk Scores

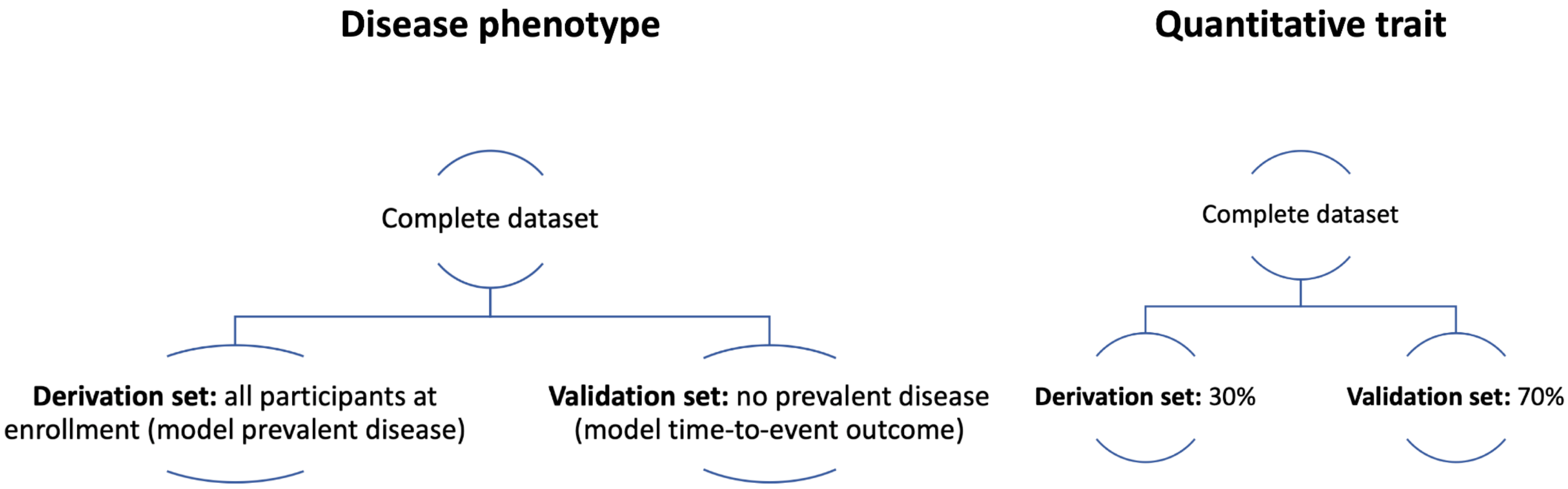


**Disease phenotype:** the derivation step used prevalent disease status as the outcome, which was defined at baseline. Prevalent cases were then removed and the scores were validated using a survival analysis with time-to-event outcome. The two steps consider different periods of risk and thus can be seen as relatively independent.

### Supplemental Figure 2. Risk prediction scores for atrial fibrillation

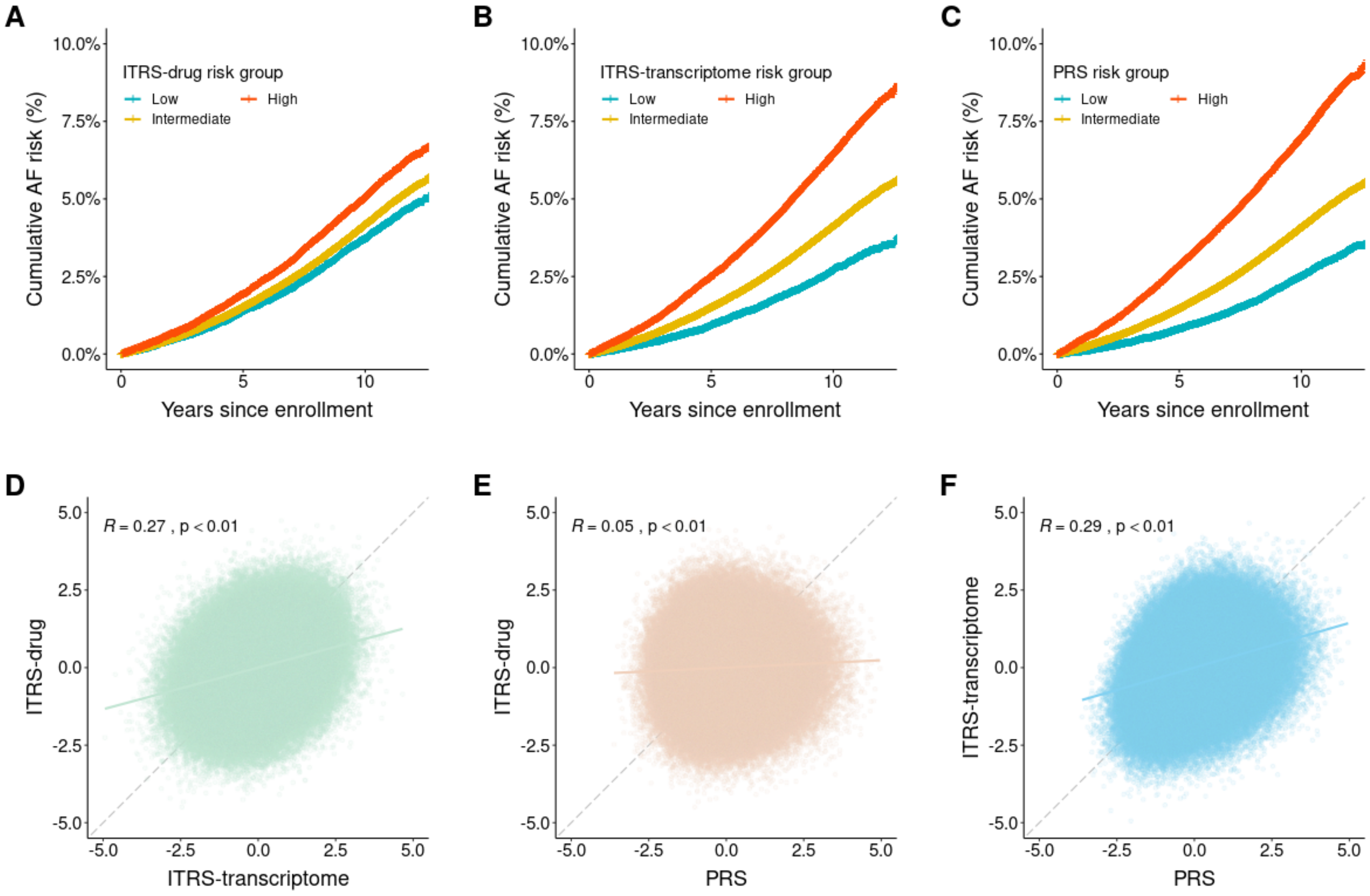

**AF:** atrial fibrillation; **ITRS-transcriptome:** imputed transcriptome risk score constructed using suggestive significant tissue-gene pairs observed in transcriptome-wide association study (TWAS); **ITRS-drug:** ITRS constructed using a subset of suggestive significant tissue-gene pairs observed in TWAS, for which the genes are actionable proteins defined in a previous study; **PRS:** genome-wide polygenic risk score. All risk scores are plotted in standardized units calculated using the entire testing sample (N=478,165). **(A)** cumulative risk of AF stratified by ITRS-drug risk groups (Low: lowest 10%, Intermediate: middle 80%, High: highest 10%); **(B)** cumulative risk of AF stratified by ITRS-transcriptome risk groups; **(C)** cumulative risk of AF stratified by PRS risk groups; **(D)** correlation between ITRS-transcriptome and ITRS-drug; ***R***: Pearson correlation coefficient; **(E)** correlation between PRS and ITRS-drug; **(F)** correlation between PRS and ITRS-transcriptome.

### Supplemental Figure 3. Risk prediction scores for type 2 diabetes

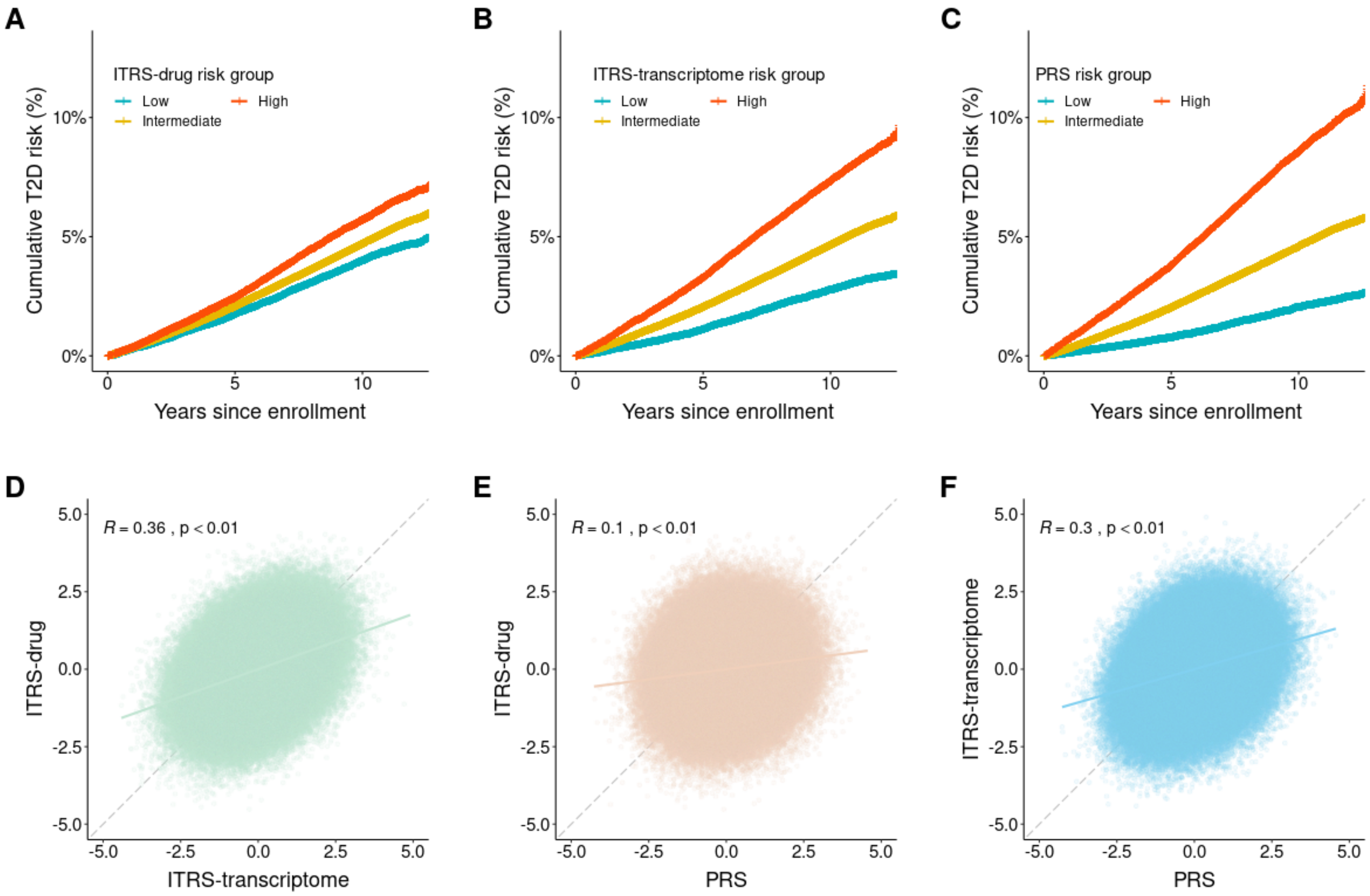

**T2D:** type 2 diabetes; **ITRS-transcriptome:** imputed transcriptome risk score constructed using suggestive significant tissue-gene pairs observed in transcriptome-wide association study (TWAS); **ITRS-drug:** ITRS constructed using a subset of suggestive significant tissue-gene pairs observed in TWAS, for which the genes are actionable proteins defined in a previous study; **PRS:** genome-wide polygenic risk score. All risk scores are plotted in standardized units calculated using the entire testing sample (N=474,481). **(A)** cumulative risk of T2D stratified by ITRS-drug risk groups (Low: lowest 10%, Intermediate: middle 80%, High: highest 10%); **(B)** cumulative risk of T2D stratified by ITRS-transcriptome risk groups; **(C)** cumulative risk of T2D stratified by PRS risk groups; **(D)** correlation between ITRS-transcriptome and ITRS-drug; ***R***: Pearson correlation coefficient; **(E)** correlation between PRS and ITRS-drug; **(F)** correlation between PRS and ITRS-transcriptome.

Supplemental Figure 4. Risk prediction scores for QTc interval on ECGs

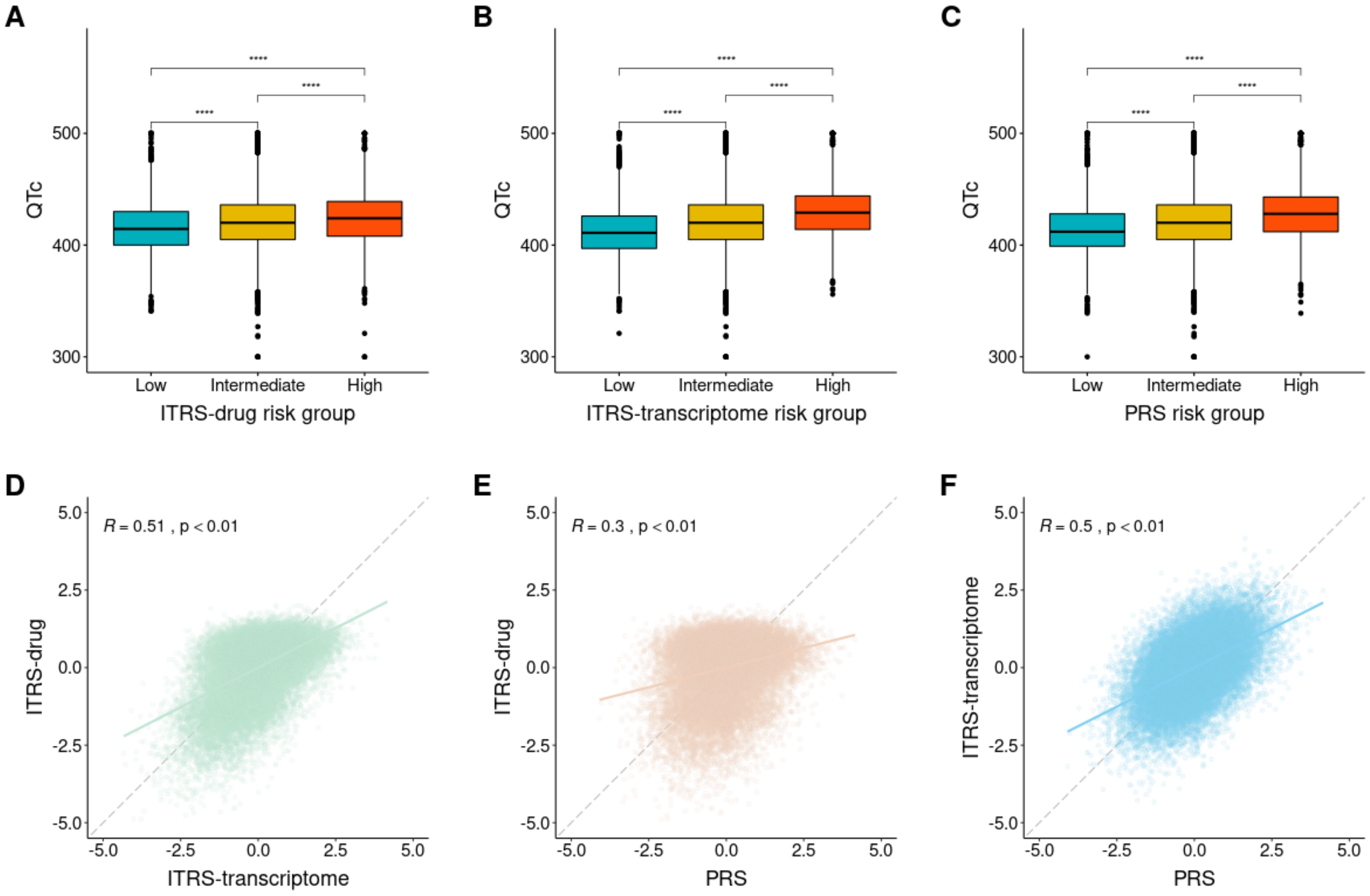

**QTc:** heart rate-corrected QT interval on electrocardiograms (ECGs); **ITRS-transcriptome:** imputed transcriptome risk score constructed using suggestive significant tissue-gene pairs observed in transcriptome-wide association study (TWAS); **ITRS-drug:** ITRS constructed using a subset of suggestive significant tissue-gene pairs observed in TWAS, for which the genes are actionable proteins defined in a previous study; **PRS:** genome-wide polygenic risk score. All risk scores are plotted in standardized units calculated using the entire testing sample (N=29,053). **(A)** distributions of QTc values across ITRS-drug risk groups (Low: lowest 10%, Intermediate: middle 80%, High: highest 10%); **(B)** distributions of QTc values across ITRS-transcriptome risk groups; **(C)** distributions of QTc values across PRS risk groups; **(D)** correlation between ITRS-transcriptome and ITRS-drug; ***R***: Pearson correlation coefficient; **(E)** correlation between PRS and ITRS-drug; **(F)** correlation between PRS and ITRS-transcriptome.

### Supplemental Figure 5. Risk prediction scores for glycated hemoglobin

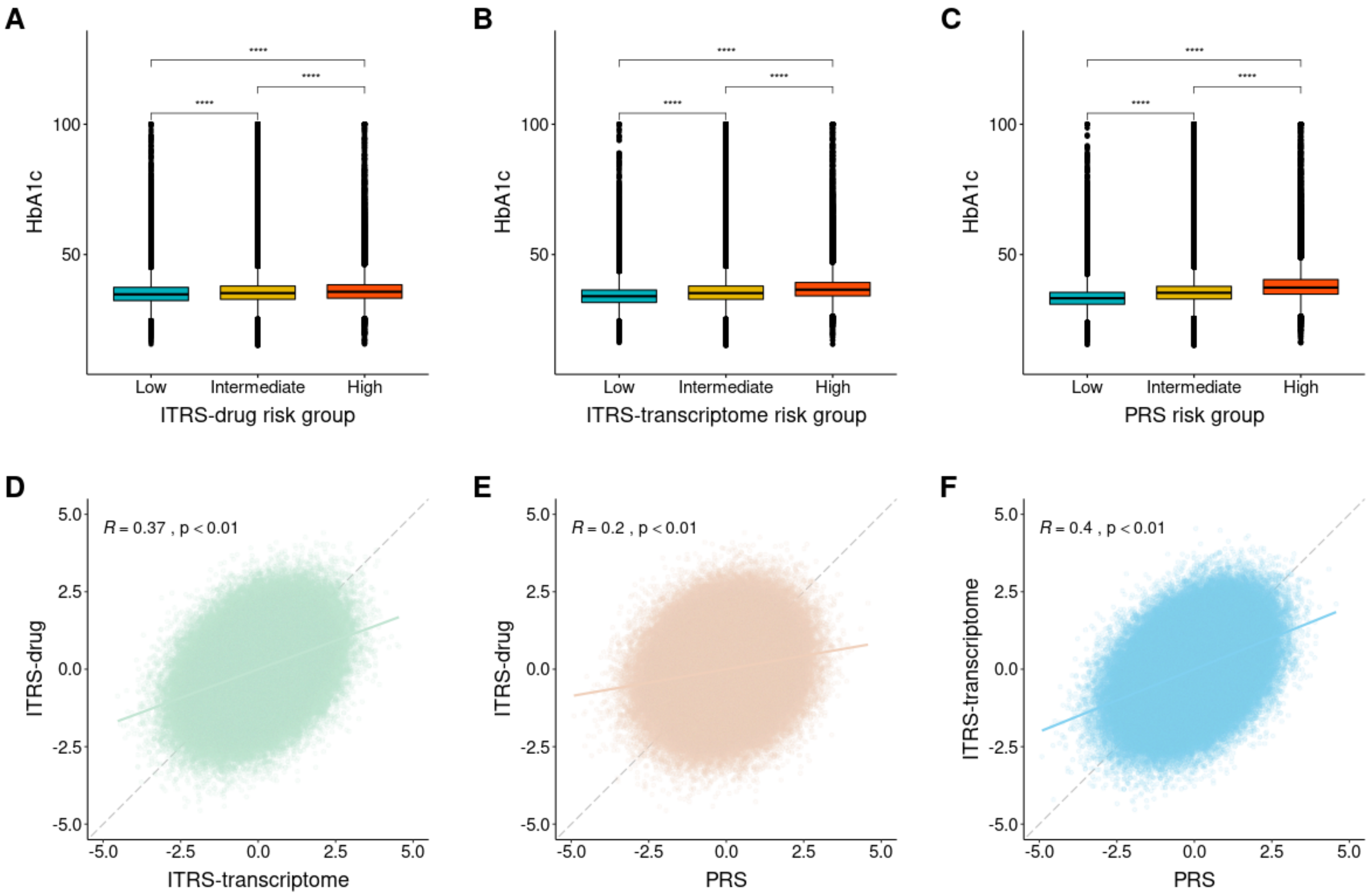

**HbA1c:** hemoglobin A1C (glycated hemoglobin); **ITRS-transcriptome:** imputed transcriptome risk score constructed using suggestive significant tissue-gene pairs observed in transcriptome-wide association study (TWAS); **ITRS-drug:** ITRS constructed using a subset of suggestive significant tissue-gene pairs observed in TWAS, for which the genes are actionable proteins defined in a previous study; **PRS:** genome-wide polygenic risk score. All risk scores are plotted in standardized units calculated using the entire testing sample (N=323,542). **(A)** distributions of HbA1c levels across ITRS-drug risk groups (Low: lowest 10%, Intermediate: middle 80%, High: highest 10%); **(B)** distributions of HbA1c levels across ITRS-transcriptome risk groups; **(C)** distributions of HbA1c levels across PRS risk groups; **(D)** correlation between ITRS-transcriptome and ITRS-drug; ***R***: Pearson correlation coefficient; **(E)** correlation between PRS and ITRS-drug; **(F)** correlation between PRS and ITRS-transcriptome.

Supplemental Figure 6. Genes representing therapeutic targets with at least one significant association identified in the whole blood phenome-wide association study

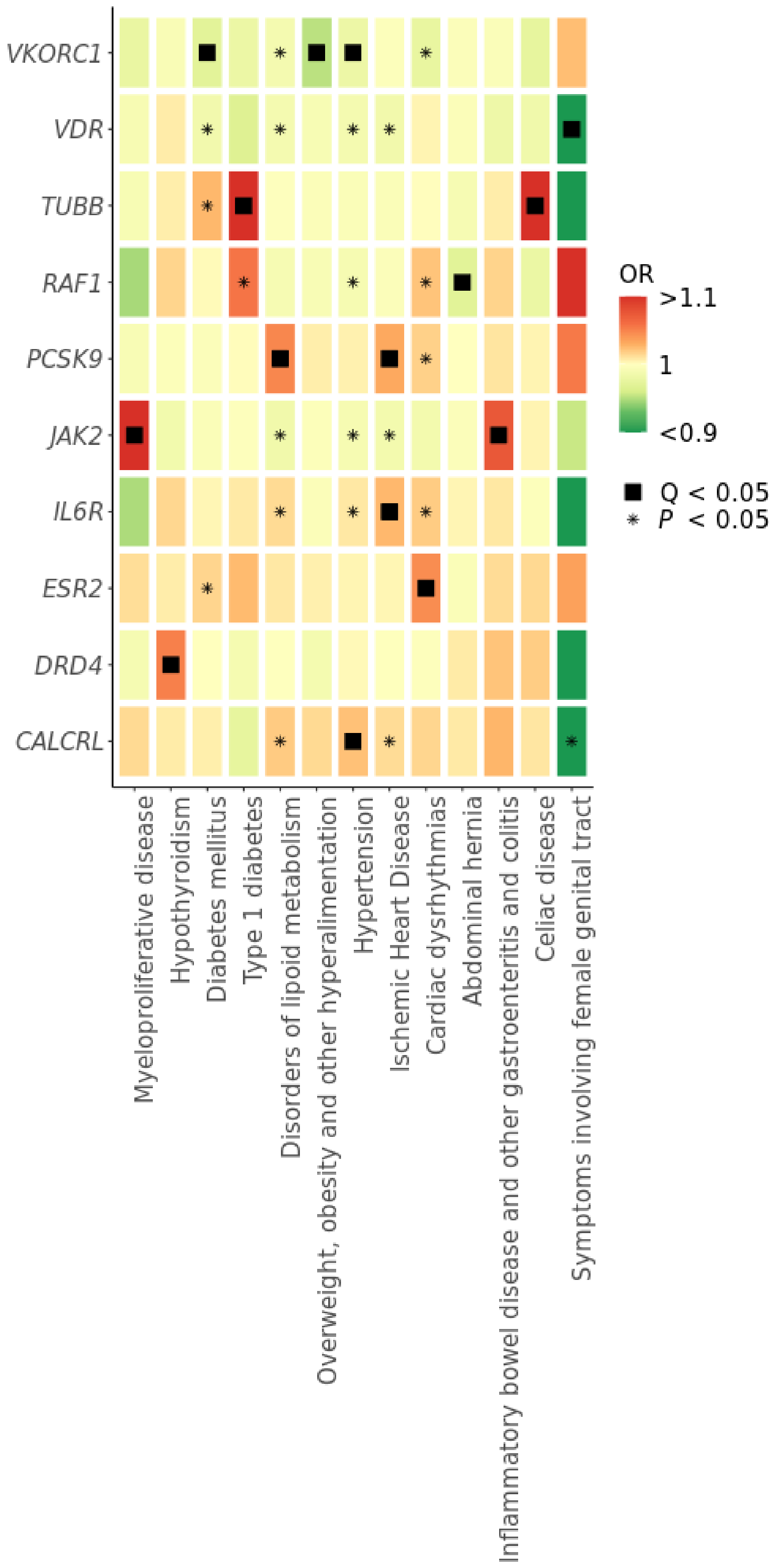
VKORC1
VDR
TUBB
RAF1
PCSK9
JAK2
IL6R
ESR2
DRD4
CALCRL
OR
>1.1
1
<0.9
Q < 0.05
P < 0.05
Myeloproliferative disease
Hypothyroidism
Diabetes mellitus
Type 1 diabetes
Disorders of lipoid metabolism
Overweight, obesity and other hyperalimentation
Hypertension
Ischemic Heart Disease
Cardiac dysrhythmias
Abdominal hernia
Inflammatory bowel disease and other gastroenteritis and colitis
Celiac disease
Symptoms involving female genital tract

Established therapeutic targets (genes) are plotted on the y-axis. Phenotypes (phecodes) are plotted on the x-axis. Colors represent the direction and strength of associations between gene expression and phecodes. Associations with a FDR adjusted *P* value < 0.05 were annotated using black square, and associations with an unadjusted *P* value < 0.05 were annotated using asterisk.
